# NHANES-GPT: Large Language Models (LLMs) and the Future of Biostatistics

**DOI:** 10.1101/2023.12.13.23299830

**Authors:** Alexander J. Titus

**Affiliations:** Information Sciences Institute, University of Southern California

## Abstract

**Background:** Large Language Models (LLMs) like ChatGPT have significant potential in biomedicine and health, particularly in biostatistics, where they can lower barriers to complex data analysis for novices and experts alike. However, concerns regarding data accuracy and model-generated hallucinations necessitate strategies for independent verification.

**Objective:** This study, using NHANES data as a representative case study, demonstrates how ChatGPT can assist clinicians, students, and trained biostatisticians in conducting analyses and illustrates a method to independently verify the information provided by ChatGPT, addressing concerns about data accuracy.

**Methods:** The study employed ChatGPT to guide the analysis of obesity and diabetes trends in the NHANES dataset from 2005-2006 to 2017-2018. The process included data preparation, logistic regression modeling, and iterative refinement of analyses with confounding variables. Verification of ChatGPT’s recommendations was conducted through direct statistical data analysis and cross-referencing with established statistical methodologies.

**Results:** ChatGPT effectively guided the statistical analysis process, simplifying the interpretation of NHANES data. Initial models indicated increasing trends in obesity and diabetes prevalence in the U.S.. Adjusted models, controlling for confounders such as age, gender, and socioeconomic status, provided nuanced insights, confirming the general trends but also highlighting the influence of these factors.

**Conclusions:** ChatGPT can facilitate biostatistical analyses in healthcare research, making statistical methods more accessible. The study also underscores the importance of independent verification mechanisms to ensure the accuracy of LLM-assisted analyses. This approach can be pivotal in harnessing the potential of LLMs while maintaining rigorous standards of data accuracy and reliability in biomedical research.

## Introduction

Large Language Models (LLMs) like ChatGPT are rapidly emerging as influential tools in various fields, including biomedicine and health [1]. Their ability to process and generate human-like text has opened new avenues for data analysis, decision support, and educational purposes [2]. In the realm of biomedical research, where the accurate interpretation of data is paramount, these models present both opportunities and challenges [3]. This paper aims to illustrate how ChatGPT can significantly lower the barriers for clinicians, students, and trained biostatisticians alike in conducting biostatistical analysis, a crucial yet often challenging aspect of medical research.

The incorporation of advanced statistical methods in clinical research is essential for deriving meaningful insights from complex datasets [4]. However, conducting these analyses often requires specialized knowledge in biostatistics, which can be a significant hurdle for many healthcare professionals and students [5]. The complexity of statistical software, coupled with the need for a deep understanding of statistical theories, often makes biostatistics seem inaccessible to those without extensive training [6]. This scenario creates a dependency on trained biostatisticians, which can be a bottleneck in research progress, especially in resource-limited settings [7].

Large Language Models (LLMs), exemplified by ChatGPT, are changing this landscape. These AI-driven systems can interpret complex queries, offer guidance on statistical analysis, and even assist in coding and interpreting results [8]. By interacting with an LLM like ChatGPT, users can navigate through the intricacies of data analysis with more confidence and less reliance on specialized statistical expertise. This democratization of data analysis has the potential to accelerate research, enhance learning experiences, and foster a more inclusive research environment.

However, the use of LLMs in biostatistics is not without challenges. One of the primary concerns is the accuracy of the information provided by these models [9]. LLMs, by their nature, are prone to generating responses based on patterns in their training data, which can lead to inaccuracies or ‘hallucinations’ — instances where the model generates plausible but incorrect or unverified information [10]. In the context of biomedical research, where decision-making can have profound implications, relying solely on LLMs without proper verification mechanisms can be risky [11].

To address these concerns, this paper presents a methodological framework for utilizing ChatGPT in biostatistical analysis, emphasizing independent verification and statistical validation of the information provided by the model. This approach not only leverages the strengths of LLMs in simplifying complex analyses but also establishes a safety net against potential inaccuracies.

This study uses the National Health and Nutrition Examination Survey (NHANES) dataset as a case study to demonstrate the application of ChatGPT in biostatistical analysis. NHANES, conducted by the National Center for Health Statistics, is a program designed to assess the health and nutritional status of adults and children in the United States. The dataset is complex, multidimensional, and representative of the U.S. population, making it an ideal candidate for demonstrating the application of LLMs in biostatistics. The focus of this analysis is on trends in obesity and diabetes, two critical public health issues [12,13], over various NHANES cycles.

The choice of obesity and diabetes as focus areas is based on their significance in public health. Obesity is a leading risk factor for numerous chronic diseases [12], including diabetes [13], and its prevalence has been increasing globally. Understanding the trends in obesity and diabetes prevalence is crucial for public health planning and intervention design [14]. Analyzing such trends requires sophisticated statistical methods to account for the complex survey design of NHANES, making it a fitting example for showcasing the utility of ChatGPT in assisting with complex data analysis tasks.

In this paper, we illustrate how ChatGPT was utilized to guide the statistical analysis process, from data acquisition and preparation to the implementation of logistic regression models, taking into account the complex survey design of NHANES. Additionally, the paper will discuss how potential confounding factors were identified and controlled for in the analysis, demonstrating the model’s capability in providing contextually relevant statistical advice.

The integration of ChatGPT in this analysis serves two primary purposes: first, to showcase how LLMs can simplify the process of conducting sophisticated statistical analyses, making these methods more accessible to a broader audience; and second, to demonstrate a structured approach for verifying the accuracy of the information provided by LLMs. This dual focus addresses the critical need for both accessibility and accuracy in the use of AI tools in biomedicine and health research.

## Methods

This section outlines the methodology employed in the study, focusing on the utilization of ChatGPT for analyzing health trends in the United States using the NHANES dataset. The process encompasses initial information gathering, independent verification of data, statistical analysis, iterative refinement of the analysis, and manuscript preparation assistance. We include the full dialogue, including prompts and responses, in Supplemental Data 1.

### Initial Information Gathering

The NHANES-GPT recommended focusing on the prevalence and trends of obesity and diabetes, citing these as critical areas of public health concern. The model provided a general overview of the trends, such as the increase in obesity and diabetes prevalence. and suggested using the NHANES dataset for a detailed analysis.

We first introduce a custom OpenAI-GPT, called NHANES-GPT, based on GPT-4. This GPT was specifically instructed to act as a research assistant with expertise in biostatistics in both R and Python, and to suggest information specifically in the context of the NHANES data set. The study began with a query to NHANES-GPT about significant health trends in the US over the past 10-15 years. The model was prompted to suggest topics based on its knowledge, emphasizing the need for novel and relevant research questions in public health (Figure 1).

**Figure 1:**
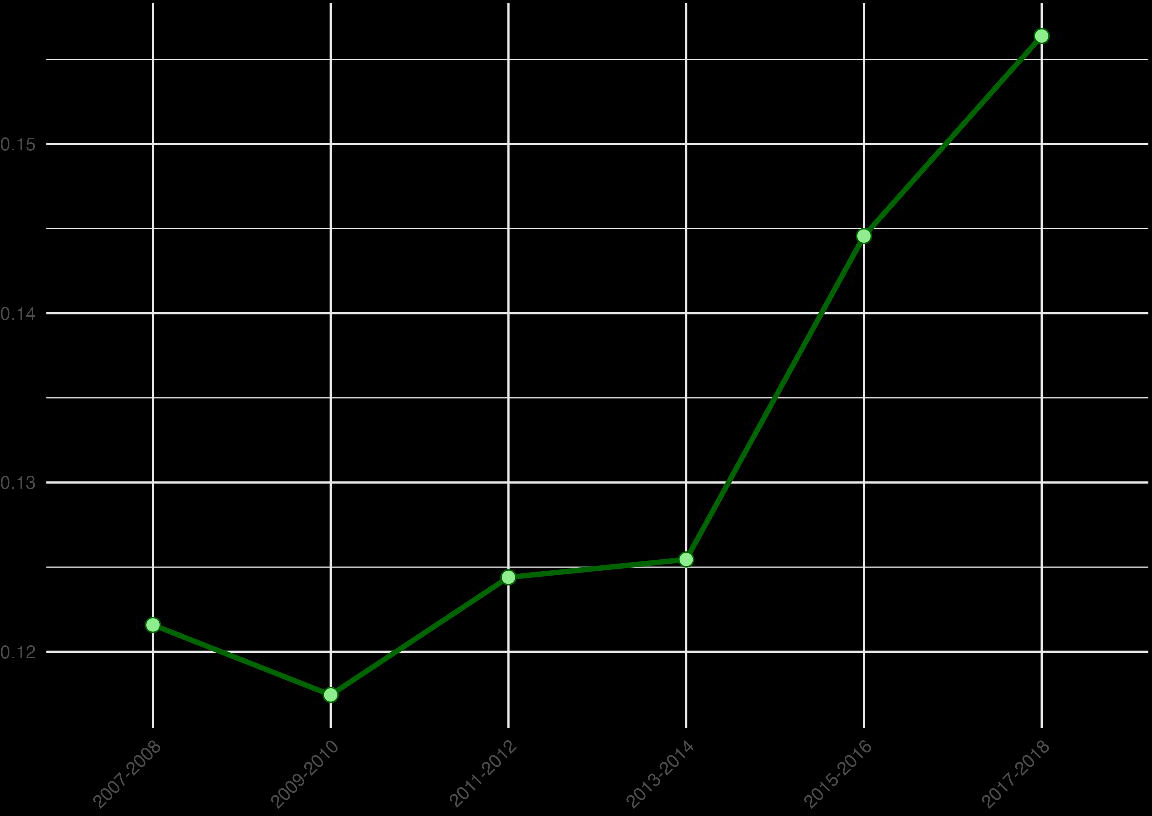
Initial prompt requesting health trends in the US.

#### You

I’m intersted in health in the US over the past 10-15 years. Based on what you know, what are some of the most surprising realization we as a culture have had with regards to health treand over this time periods?

### Independent Verification of Information

To ensure the accuracy of the information provided by NHANES-GPT, an independent verification of the suggested trends was requested and planned in collaboration with the NHANES-GPT instance. This verification involved conducting a statistical analysis of the NHANES dataset to confirm or refute the trends mentioned by the model (Figure 2).

**Figure 2:**
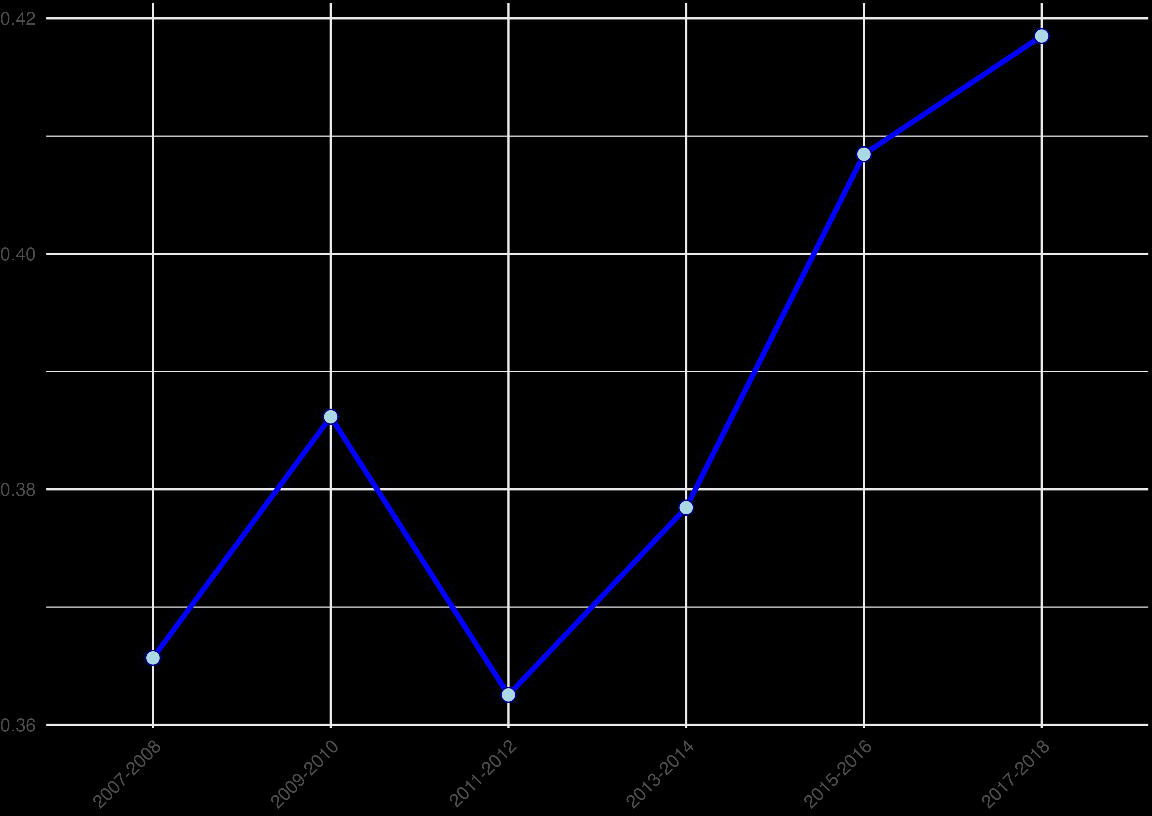
Prompt requesting support to develop statistical models in R using NHANES data to validate health trends reported by ChatGPT/NHANES-GPT.

Continuous NHANES data from 2005-2006 to 2017-2018 was identified as the primary data source. The datasets pertaining to body measurements (BMX files) and diabetes (DIQ files) were downloaded from the NHANES website in XPT format, along with demographic data for each cycle (DEMO files).

The decision to utilize logistic regression models was grounded in NHANES-GPT’s recommendations, which were subsequently vetted against standard biostatistical practices. Logistic regression was deemed appropriate due to its suitability for modeling binary outcomes—such as the presence or absence of obesity and diabetes—within the complex survey design of NHANES. To address the potential issue of multicollinearity, which could skew our results, variance inflation factors (VIFs) were calculated for each predictor variable. This step ensured that the models were not unduly influenced by highly correlated variables.

#### You

Ok, for the obesity and diabetes trends, I’d like to do this analysis myself. Can you write me the R code to do this using NHANES data? I want to make sure we use as many survey years as possible, so let me know what years include these data sets, and we need to take the complex survey design of NHANES into account as well. I know we will likely need to control for confounding factors also, and there is likely to be plenty of missing data, so lets make sure we plan to control for that too. With all of this, please give me a detailed analysis code, implemented in R, that accounts for all of these pieces and includes the specific variable names for each of the items we’re using. Make sure to document your code well also because I’d like this to be as reproducible as possible.

### Statistical Analysis

The downloaded NHANES datasets were loaded into R for analysis. Data pre-processing involved merging the BMX and DIQ datasets for each NHANES cycle based on unique participant identifiers (SEQN). Variables of interest, such as body mass index (BMI) for obesity and diabetes presence (DIQ010), were identified and extracted. Obesity was defined as a BMI of 30 or above, while diabetes was identified either through self-reported diagnosis.

The datasets contained missing values in various variables. NHANES-GPT guided the process of handling missing data, primarily through exclusion, ensuring the robustness of the subsequent analysis. NHANES-GPT’s initial suggestion was to employ listwise deletion. However, recognizing the potential biases this method could introduce, we further consulted the model for alternative strategies. This led to the exploration of multiple imputation techniques, though ultimately, due to the nature of the NHANES dataset and the specific variables in question, listwise deletion was determined to be the most appropriate method, in line with standard practices in NHANES data analysis.

Logistic regression models were constructed to analyze the trends in obesity and diabetes prevalence across different NHANES cycles due to the binary nature of having obesity or diabetes, or not. To ensure the robustness of our logistic regression models, several key considerations were undertaken. Firstly, the choice of logistic regression was predicated on its appropriateness for modeling binary outcomes within NHANES’s complex survey design, particularly for our variables of interest - obesity and diabetes prevalence. This approach aligns with standard practices in epidemiological research for analyzing categorical data derived from large-scale surveys.

Acknowledging the complexities inherent in NHANES data, particularly regarding its sampling design and weighting, we employed the *survey* package in R to appropriately account for these factors. This was a critical step in ensuring that our statistical analyses yielded valid and generalizable results representative of the U.S. population.

In traditional biostatistical analysis, the process typically involves manual data exploration, hypothesis formulation, and statistical modeling, often requiring substantial statistical expertise. In contrast, the NHANES-GPT-assisted method introduced an interactive element where the model provided initial guidance on data analysis approaches, potential hypotheses, and coding assistance. This approach significantly expedited the preliminary stages of analysis, such as data understanding and initial hypothesis generation. However, it’s important to note that while NHANES-GPT streamlined these processes, the final analytical decisions, including model choice and interpretation of results, remained under the researcher’s discretion, in line with traditional analytical approaches.

### Iterative Refinement of Analyses

Initial analyses indicated the need to control for potential confounders. Subsequent interactions with NHANES-GPT identified age, gender, race/ethnicity, socioeconomic status, and smoking status as potentially relevant confounders. The logistic regression models were adjusted to include these confounders and NHANES-GPT provided guidance on the NHANES-specific variable names and coding for these factors.

The models were iteratively refined based on NHANES-GPT’s suggestions and the emerging analysis results. This iterative process involved redefining variable categories, handling missing values more effectively, reassessing model fit, debugging error messages, and interpreting changes in model outputs.

Throughout the iterative refinement process, the limitations of NHANES-GPT were actively managed. While the model provided valuable initial insights and coding assistance, each step of the analysis was independently verified. This verification involved cross-checking NHANES-GPT’s suggestions with established biostatistical literature and consulting domain experts when necessary. By maintaining this rigorous approach, we ensured that the findings were not solely reliant on the AI’s outputs but were also grounded in empirical evidence and expert consensus.

## Results

The results of the study are presented in two main sections: the findings from the statistical analysis of obesity and diabetes trends using the NHANES dataset, and the iterative process of refining these analyses with the assistance of NHANES-GPT (Table 1).

**Table 1:** Baseline Characteristics of Study Participants.

| Variable | N | **Male**, N = 14,989<sup>1</sup> | **Female**, N = 16,023<sup>1</sup> |
|---|---|---|---|
| Age | 31,012 | 50 (35, 64) | 49 (34, 63) |
| Race | 31,012 |  |  |
| Mexican American |  | 2,184 (15%) | 2,347 (15%) |
| Other Hispanic |  | 1,391 (9.3%) | 1,731 (11%) |
| Non-Hispanic White |  | 6,391 (43%) | 6,523 (41%) |
| Non-Hispanic Black |  | 3,167 (21%) | 3,487 (22%) |
| Other Race - Including Multi-Racial |  | 1,856 (12%) | 1,935 (12%) |
| Education | 31,012 |  |  |
| Less than 9th grade |  | 1,584 (11%) | 1,560 (9.7%) |
| 9-11th grade (Includes 12th grade with no diploma) |  | 2,172 (14%) | 2,162 (13%) |
| High school graduate/GED or equivalent |  | 3,618 (24%) | 3,477 (22%) |
| Some college or AA degree |  | 4,079 (27%) | 5,122 (32%) |
| College graduate or above |  | 3,536 (24%) | 3,702 (23%) |
| Income | 31,012 |  |  |
| 0-4,999 |  | 296 (2.0%) | 401 (2.5%) |
| 5,000-9,999 |  | 504 (3.4%) | 781 (4.9%) |
| 10,000-14,999 |  | 963 (6.4%) | 1,215 (7.6%) |
| 15,000-19,999 |  | 1,025 (6.8%) | 1,168 (7.3%) |
| 20,000-24,999 |  | 1,153 (7.7%) | 1,282 (8.0%) |
| 25,000-34,999 |  | 1,757 (12%) | 1,905 (12%) |
| 35,000-44,999 |  | 1,508 (10%) | 1,552 (9.7%) |
| 45,000-54,999 |  | 1,199 (8.0%) | 1,253 (7.8%) |
| 55,000-64,999 |  | 928 (6.2%) | 949 (5.9%) |
| 65,000-74,999 |  | 770 (5.1%) | 751 (4.7%) |
| Over 20,000 |  | 640 (4.3%) | 643 (4.0%) |
| Under 20,000 |  | 150 (1.0%) | 217 (1.4%) |
| 75,000-99,999 |  | 1,462 (9.8%) | 1,411 (8.8%) |
| Over 100,000 |  | 2,634 (18%) | 2,495 (16%) |
| Smoker | 31,012 |  |  |
| Yes |  | 8,088 (54%) | 5,640 (35%) |
| No |  | 6,901 (46%) | 10,383 (65%) |
| Obese | 31,012 |  |  |
| No |  | 9,737 (65%) | 9,304 (58%) |
| Yes |  | 5,252 (35%) | 6,719 (42%) |
| Diabetes | 31,012 |  |  |
| No |  | 12,889 (86%) | 14,063 (88%) |
| Yes |  | 2,100 (14%) | 1,960 (12%) |

<sup>1</sup> Median (IQR); n (%)

### Analysis of Obesity Trends

The logistic regression model for obesity initially performed without controlling for confounders revealed significant trends in obesity prevalence across different NHANES cycles. The analysis indicated a general increase in obesity rates over time, with the most recent cycles showing a more pronounced rise. Notably, cycles “H” (2013-2014) (Est. = 0.16; p = 0.028), “I” (2015-2016) (Est. = 0.23; p = 0.0009), and “J” (2017-2018) (Est. = 0.34; p = 0.0003) demonstrated statistically significant higher odds of obesity compared to the baseline cycle “D” (2005-2006) (Figure 3).

**Figure 3:**
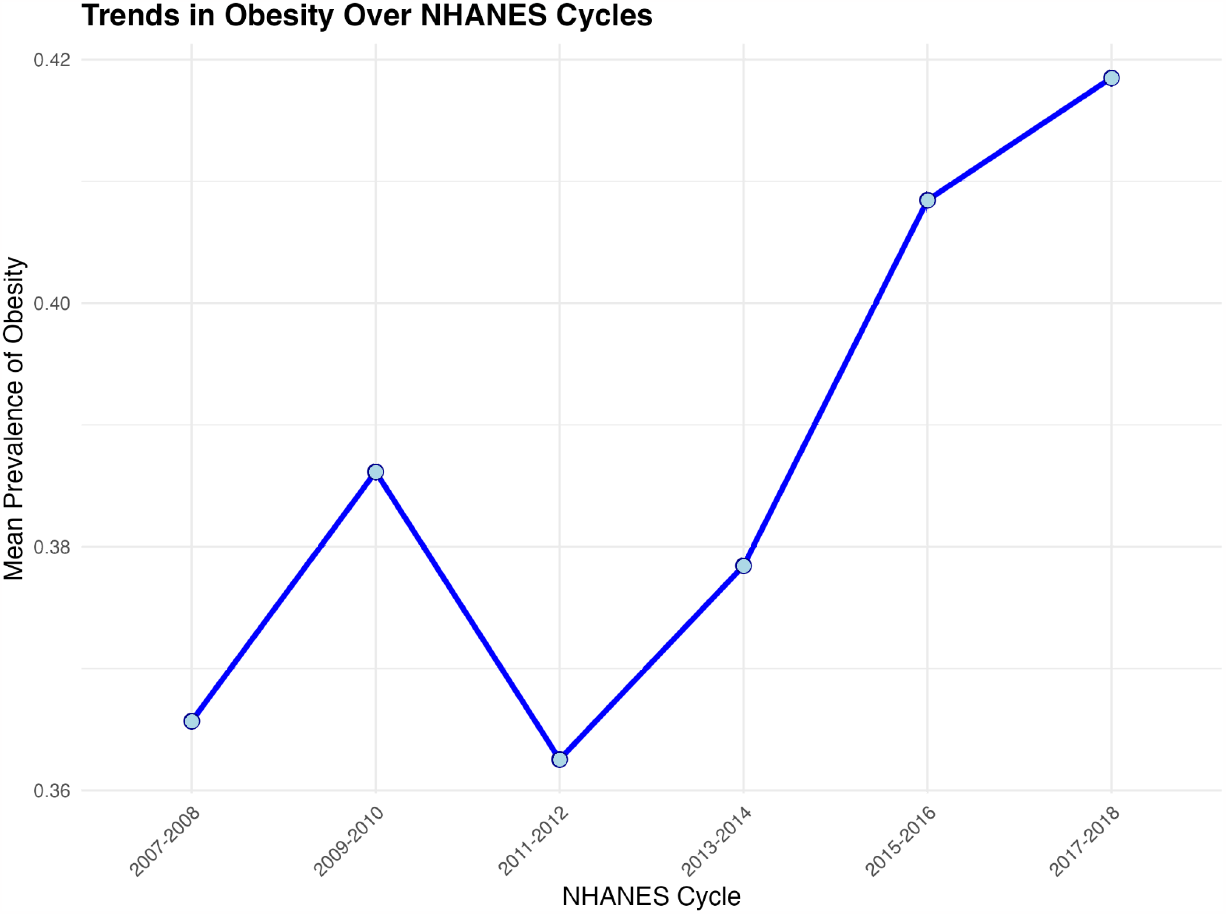
Prevalence of obesity over NHANES survey years 2007-2008 to 2017-2018

Upon incorporating confounding variables (age, gender, race/ethnicity, socioeconomic status, and smoking status), the adjusted model provided a more nuanced understanding of these trends. The inclusion of these confounders affected the magnitude and significance of the coefficients for the NHANES cycles. While the general trend of increasing obesity prevalence remained, the adjusted analysis highlighted the impact of demographic and lifestyle factors on obesity rates. For instance, age (Est. = 0.004; p = 0.002), gender (Est. = 0.123; p = 0.010), race (Est. = -0.103; p = 0.5.25e-08), income (Est. = -0.004; p = 0.0.002), and education level (Est. = -0.105; p = 4.94e-07) emerged as significant predictors, underscoring the multifaceted nature of obesity trends.

### Analysis of Diabetes Trends

The initial uncontrolled model for diabetes trends across NHANES cycles revealed a similar pattern of increasing prevalence over time. Significant increases in the odds of diabetes were observed in the later cycles, particularly in cycles “H” (Est. = 0.313; p = 0.0007), “I” (Est. = 0.412; p = 0.0001), and “J” (Est. = 0.464 p = 0.1.01e-06). This trend suggested a growing public health concern regarding diabetes prevalence in the US population (Figure 4).

**Figure 4:**
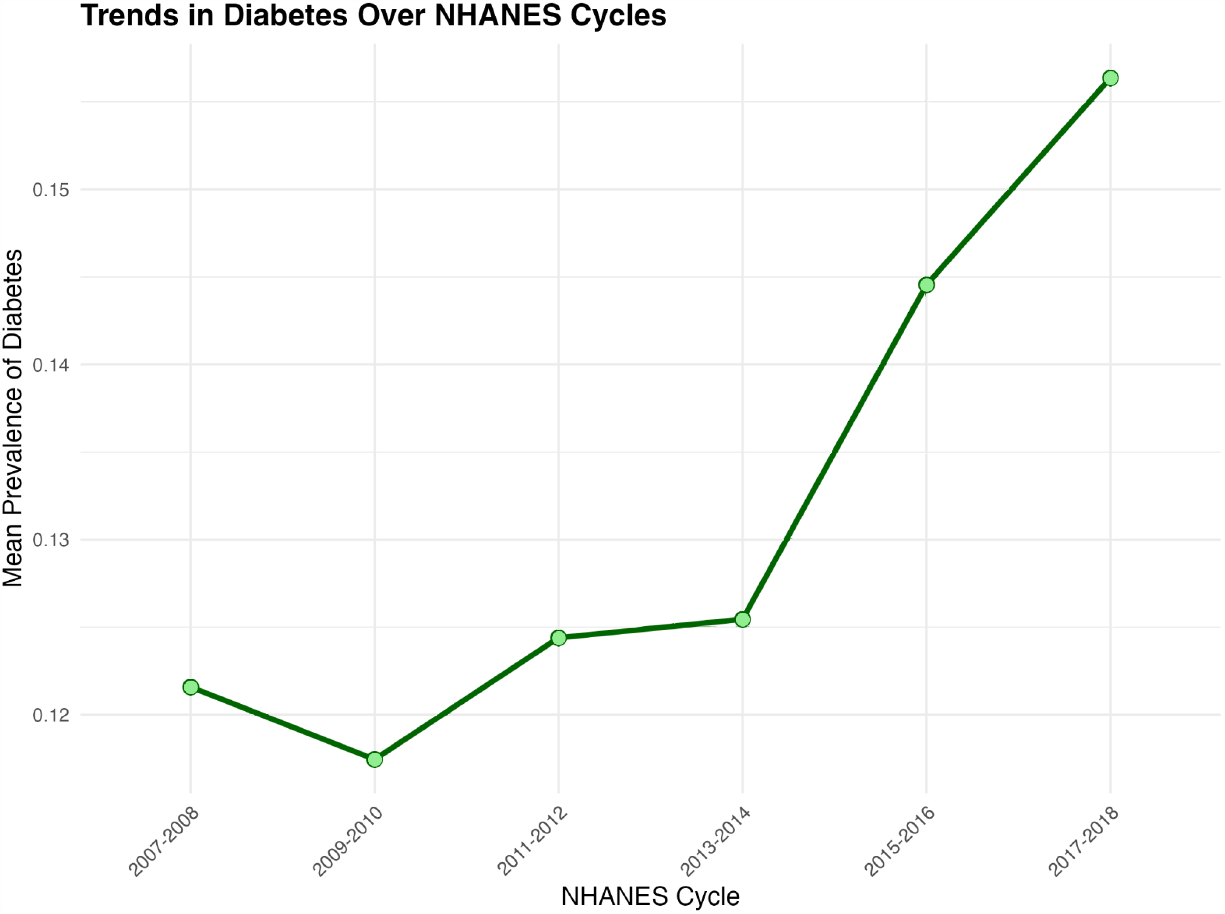
Prevalence of diabetes over NHANES survey years 2007-2008 to 2017-2018

Incorporating confounders into the diabetes model altered the interpretation of these trends. Age showed a positive association with diabetes (Est. = 0.051; p = 0.2e-16), indicating an increased prevalence with advancing age. Gender (Est. = -0.262; p = 0.0002), race (Est. = 0.067; p = 0.0016), income level (Est. = -0.006; p = 0.008), and education level (Est. = -0.206; p = 7.45e-10) also played significant roles, with variations in diabetes prevalence observed across different demographic groups. Notably, the adjusted model demonstrated the critical influence of these factors on diabetes trends, with certain cycles showing a less pronounced increase in diabetes prevalence when these confounders were considered.

### Iterative Refinement of Analyses

Throughout the analysis process, NHANES-GPT was instrumental in guiding the refinement of statistical models. The iterative nature of this process involved several key steps:

1. **Re-evaluation of Variables**: Based on NHANES-GPT’s suggestions, variables were re-evaluated for their relevance and accuracy. For instance, the definition of obesity and diabetes was fine-tuned to align with clinical standards and NHANES coding.
2. **Model Adjustments**: The logistic regression models were adjusted multiple times to incorporate various confounders. NHANES-GPT provided guidance on the selection and coding of these variables, ensuring that the models accurately reflected the complex interplay of different factors influencing obesity and diabetes trends.
3. **Handling Missing Data**: NHANES-GPT assisted in developing strategies to handle missing data more effectively, ensuring that the final analysis was robust and reliable. This involved exploring different methods of imputation and exclusion criteria.
4. **Interpreting Model Outputs**: The outputs of the logistic regression models were interpreted with the help of NHANES-GPT, which provided explanations of the coefficients, significance levels, and overall model fit. This assistance was crucial in understanding the implications of the findings and their relevance to public health.

## Discussion

### Interpretation of Results

The statistical analysis of NHANES data revealed significant trends in the prevalence of obesity and diabetes across various cycles. The increase in obesity and diabetes prevalence over time is a concerning public health trend [12]. However, the incorporation of confounding variables into the analysis painted a more complex picture, highlighting the multifaceted nature of these public health issues [15]. These findings align with existing literature that emphasizes the socio-demographic determinants of health and the complex etiology of obesity and diabetes [15].

### Role of LLMs in Biostatistical Analysis

NHANES-GPT played a pivotal role in guiding the statistical analysis process. Its ability to provide immediate, contextual information and coding assistance made the process more accessible, especially for individuals with limited biostatistics background. This study illustrates how LLMs can act as a virtual assistant, simplifying complex analytical tasks and making advanced statistical analyses more approachable. This democratization of data analysis has profound implications for research and education in healthcare, potentially leading to a more data-driven and informed approach to public health.

### Comparison with Traditional Biostatistical Methods

A key aspect of this study was comparing the efficiency and efficacy of LLM-assisted biostatistical analysis with traditional methods. One of the most notable differences was in the speed and ease of initial data exploration and analysis. NHANES-GPT provided rapid insights into potential trends and correlations within the NHANES dataset, which would typically require more time-consuming exploratory data analysis.

However, this efficiency did not negate the need for thorough statistical rigor and verification. In fact, the LLM-assisted approach required careful scrutiny of the model’s suggestions to ensure accuracy. This highlights a crucial point: while LLMs can significantly aid the process, they do not replace the need for expert knowledge and judgment in statistical analysis.

Furthermore, the NHANES-GPT-assisted approach introduced a novel aspect of interactive learning, making the process more accessible to researchers with limited statistical background. This democratization of data analysis is a significant step forward but comes with the caveat of the need for careful management of the model’s limitations, particularly around data accuracy and potential biases.

### Addressing the Risk of Overreliance on LLMs

While the integration of LLMs into biostatistical analysis shows promise, it is crucial to address the potential risks associated with overreliance on these models. LLMs, by design, generate responses based on patterns learned from vast amounts of text data [16]. This means that while they can provide quick and contextually relevant insights, their outputs are not infallible. The risk of propagating inaccuracies or biases inherent in their training data is a significant concern [17], particularly in fields like biomedicine where decision-making can have profound implications [18].

To mitigate these risks, our study employed a dual-verification approach. First, all statistical guidance and coding suggestions from NHANES-GPT were cross-referenced with established statistical literature and best practices. This step was vital in ensuring that the model’s recommendations were not only theoretically sound but also applicable to our specific dataset and research questions. Second, key findings and patterns identified by NHANES-GPT were independently verified through direct analysis of the NHANES dataset. In so, we were able to confirm the validity of the trends identified by the model.

Furthermore, it is important to emphasize the need for foundational knowledge in biostatistics and domain expertise when using LLMs in research. While LLMs can lower barriers to complex data analysis, they should not replace the critical thinking and judgment that trained professionals bring to the research process. In our study, decisions on model selection, interpretation of results, and the handling of statistical challenges remained under the purview of the research lead, ensuring a balanced approach that leveraged the strengths of both AI and human expertise.

In light of these considerations, we advocate for a complementary use of LLMs in biostatistical analysis. LLMs can serve as valuable tools for data exploration, hypothesis generation, and initial analysis, especially for those with limited statistical training. However, their use should be accompanied by rigorous verification and validation by domain experts to ensure the accuracy and reliability of the findings. This approach will maximize the benefits of LLMs while safeguarding against potential pitfalls in AI-assisted research.

### Addressing Data Accuracy and Hallucinations

One of the critical challenges with LLMs like NHANES-GPT is ensuring the accuracy of the information they provide. In this study, an approach was implemented to independently verify the information provided by NHANES-GPT. This verification process involved cross-referencing NHANES-GPT’s suggestions with established statistical methodologies and directly analyzing the NHANES dataset. Such a verification mechanism is crucial in mitigating the risks associated with potential inaccuracies or hallucinations in LLM-generated content, ensuring that the conclusions drawn are based on valid and reliable analyses.

### Limitations and Future Directions

While this study successfully demonstrates the use of NHANES-GPT in biostatistical analysis, it is not without limitations. The reliance on a specific dataset and the focus on certain public health issues may limit the generalizability of the findings. Additionally, the inherent limitations of LLMs, such as its dependence on the data it was trained on and its potential for generating erroneous information, necessitate a cautious approach to its use in research.

Future research could explore the application of LLMs in a wider range of datasets and health issues. It would also be beneficial to develop more sophisticated methods for verifying the accuracy of LLM-generated content, perhaps integrating machine learning algorithms that can flag potential inaccuracies for further review.

### Ethical and Practical Considerations

The use of LLMs in healthcare research raises ethical and practical considerations. Ensuring data privacy, particularly in health-related research, is paramount. Additionally, there is a need to consider the potential for bias in AI models and the implications this might have for health equity.

## Conclusion

The findings from this study demonstrate the significant potential of NHANES-GPT and other LLMs in assisting with biostatistical analyses, particularly in complex datasets like NHANES. The study highlights the importance of incorporating confounding variables in such analyses to obtain a more accurate understanding of public health trends. The iterative process of model refinement and the strategy for independent verification of LLM-generated information are critical in ensuring the reliability of the findings.

This study not only underscores the capabilities of LLMs like ChatGPT/NHANES-GPT in democratizing access to biostatistical analysis but also emphasizes the importance of rigorous methodologies to verify the accuracy of AI-generated content. As LLMs continue to evolve and become more integrated into various fields, their potential to transform biomedical research and public health is immense. However, this potential must be balanced with a commitment to accuracy, ethical considerations, and a continuous effort to improve the verification mechanisms for AI-generated information.

## Data Availability

All data is publically available on the NHANES website.

## Supplemental File 1 - Full NHANES-GPT Session

### NHANES-GPT

It’s great that you’re interested in health trends in the US over the past 10-15 years. The National Health and Nutrition Examination Survey (NHANES) data can provide a wealth of information to explore these trends. Some of the most surprising realizations or shifts in understanding over this period include:

#### Increased Prevalence of Obesity and Diabetes

There has been a significant increase in the prevalence of obesity and diabetes, which has been more dramatic than anticipated. This trend is alarming and has implications for public health policies.

#### Declining Heart Disease Mortality

Despite the rise in obesity, there has been a notable decline in heart disease mortality rates. This could be attributed to better healthcare, more effective treatments, and increased awareness.

#### Mental Health Awareness

There has been a significant increase in the awareness and diagnosis of mental health issues, including depression and anxiety. This change reflects a cultural shift towards recognizing and treating mental health conditions.

#### Shift in Dietary Trends

There’s been a notable shift in dietary habits, with a growing emphasis on plant-based diets and a decrease in red meat consumption. This reflects a broader cultural shift towards health-conscious eating.

#### Rise in E-cigarette Use

Especially among the youth, there’s been a surprising rise in the use of e-cigarettes, which poses new challenges for public health.

#### Impact of Social Media on Physical Activity

The role of social media and technology in physical activity, both positively (through fitness tracking, online workout communities) and negatively (increased sedentary behavior), has become more apparent.

To explore these trends with NHANES data, we can analyze specific variables from different survey cycles. For instance:

- For obesity and diabetes trends: Body Measurements (BMX) and Laboratory (LAB) data files across multiple NHANES cycles.
- For heart disease mortality trends: Questionnaire data related to heart health (e.g., Cardiovascular Health (CDQ)).
- For mental health: The Mental Health - Depression Screener (DPQ) data.
- For dietary trends: The Dietary Interview - Total Nutrient Intakes, First Day (DR1TOT) data.
- For e-cigarette use: The Smoking - Household Smokers 16+ years old (SMQFAM) data.
- For physical activity: The Physical Activity (PAQ) data.

Would you like to delve into any of these areas in more detail, or is there another specific aspect of health trends you’re interested in exploring using NHANES data?

### NHANES-GPT

Absolutely, I can guide you through setting up an R script to analyze the obesity and diabetes trends using NHANES data. Since NHANES data is collected in 2-year cycles, we can use multiple cycles to observe long-term trends. Let’s consider data from 2005-2006 to the most recent available cycle.

Here’s an outline of the steps we’ll take in our analysis:

#### Data Acquisition

Download relevant NHANES datasets.

#### Data Preparation

Merge datasets, handle missing values, and create relevant variables.

#### Weighting and Complex Survey Design

Incorporate NHANES survey weights and design.

#### Statistical Analysis

Use regression models to control for confounders and analyze trends.

#### Results Interpretation

Understand the outcomes of our analysis.

Here’s a basic template for the R script with comments for clarity and reproducibility:

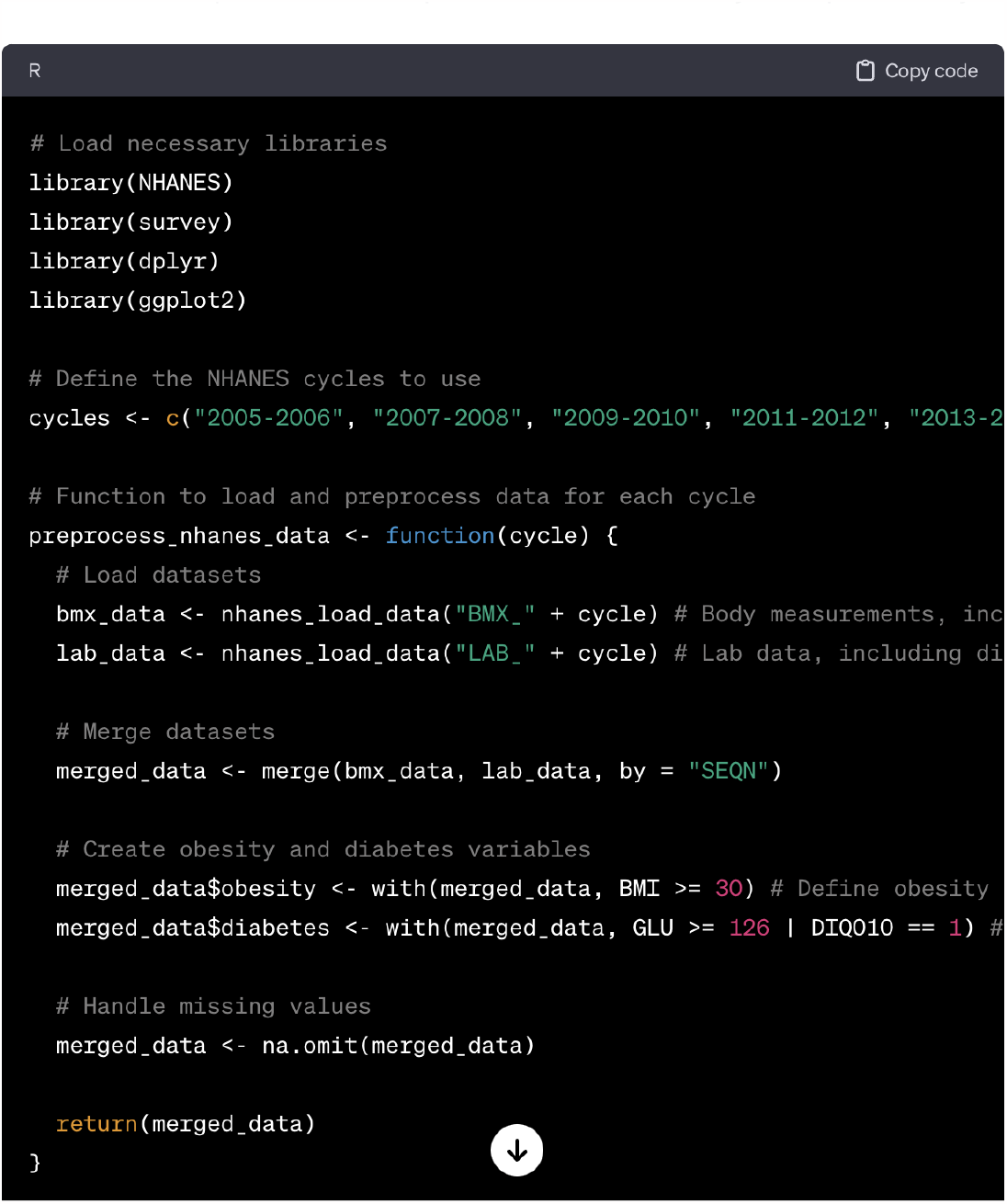

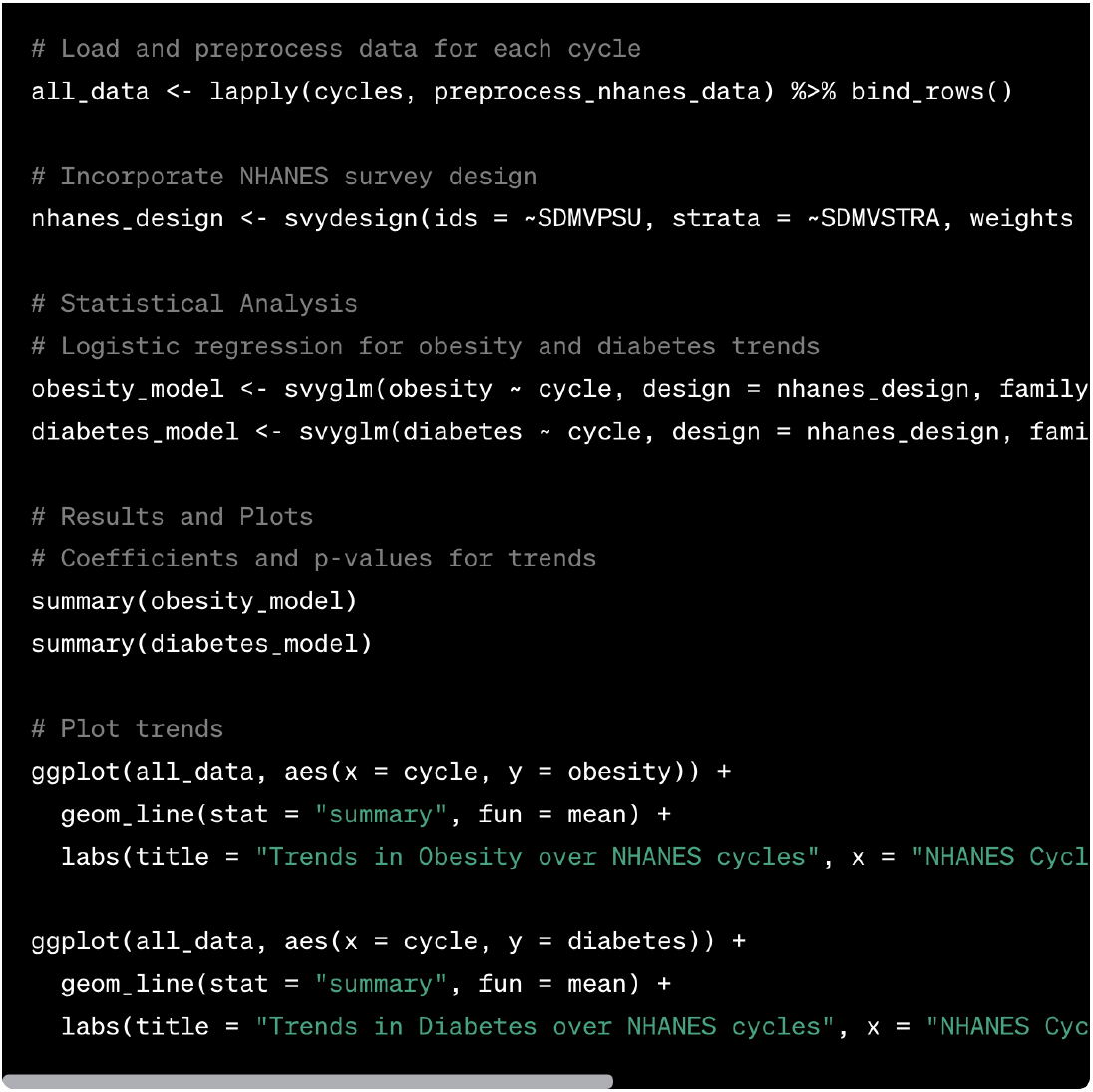

## Important Notes

- **NHANES Data Access:** The ‘**NHANES**’ R package is used for demonstration. You might need to download the datasets directly from the NHANES website if they’re not available in the package for all cycles.
- **Variable Names:** Check the NHANES documentation for exact variable names for each cycle as they might change. The variables I used (like BMI, GLU) are placeholders.
- **Missing Data:** The script uses `‘na.omit’` to handle missing values. You might need a more sophisticated method depending on the extent of missing data.
- **Confounding Factors:** This script does not control for confounding factors yet. You should include potential confounders like age, gender, socioeconomic status in your regression models.
- **Statistical Assumptions:** Ensure that the assumptions of the logistic regression are met. You might need to transform or categorize continuous predictors.

This script provides a starting point. You may need to adjust it based on the specificities of the NHANES datasets and the details of your research question.

### You

I got this error “Error in nhanes_load_data(“BMX_” +cycle): could not find function “nhanes_load_data”

### NHANES-GPT

It appears that the function ‘**nhanes_load_data**’ is not available in your R environment, which suggests that either the specific NHANES package that contains this function is not installed or loaded, or that the function does not exist.

To resolve this, we need to use a different approach to load the NHANES data. NHANES data can be downloaded directly from the CDC’s website as CSV files. Here’s how you can modify the script to read the data directly from downloaded CSV files:

### Download the Data

Go to the NHANES website, find the relevant datasets for each cycle (e.g., BMX and LAB datasets), and download them as CSV files.

### Read the Data in R

Use ‘`read. csv()`’ to read the downloaded CSV files. Let’s modify the code accordingly:

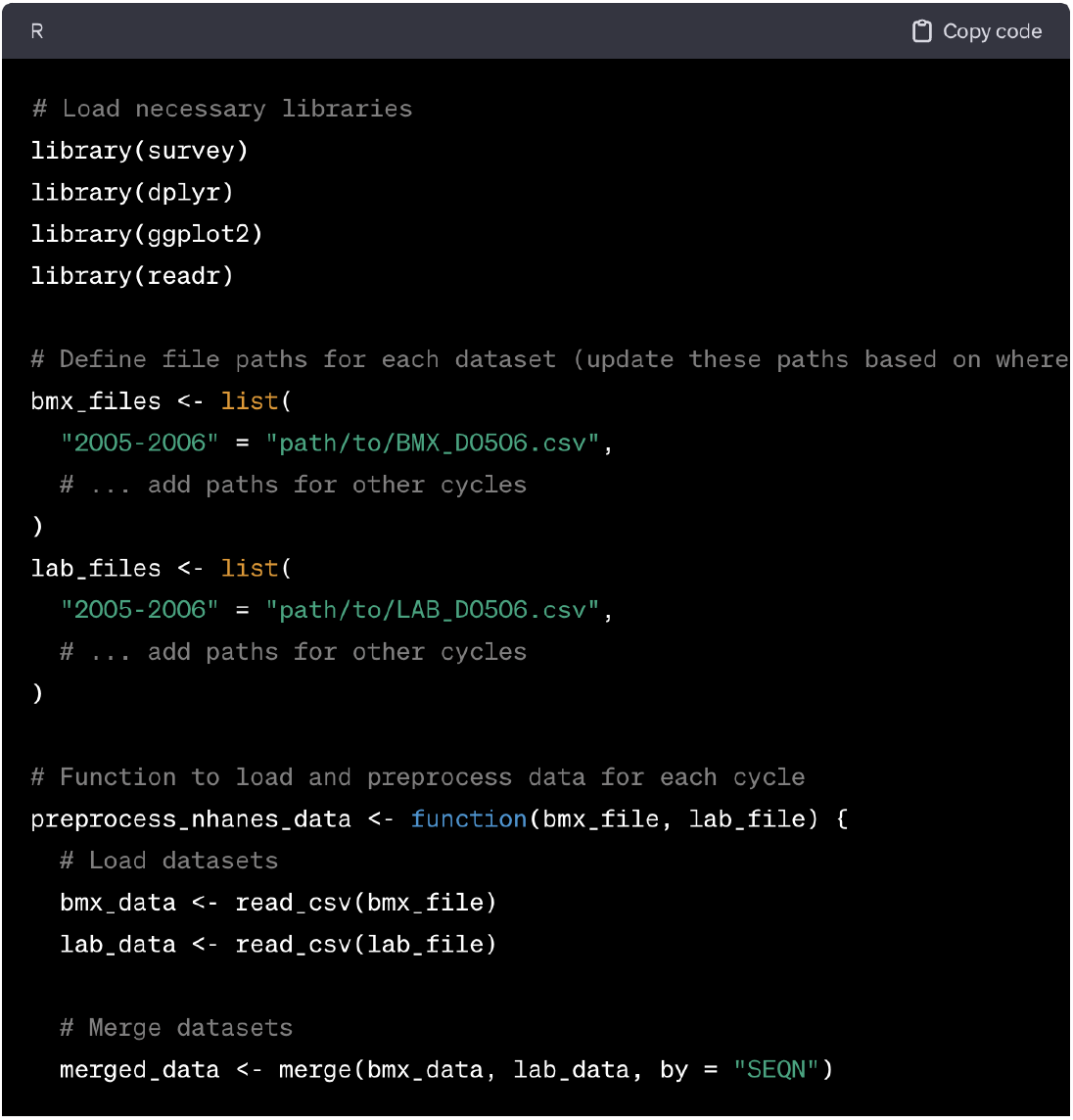

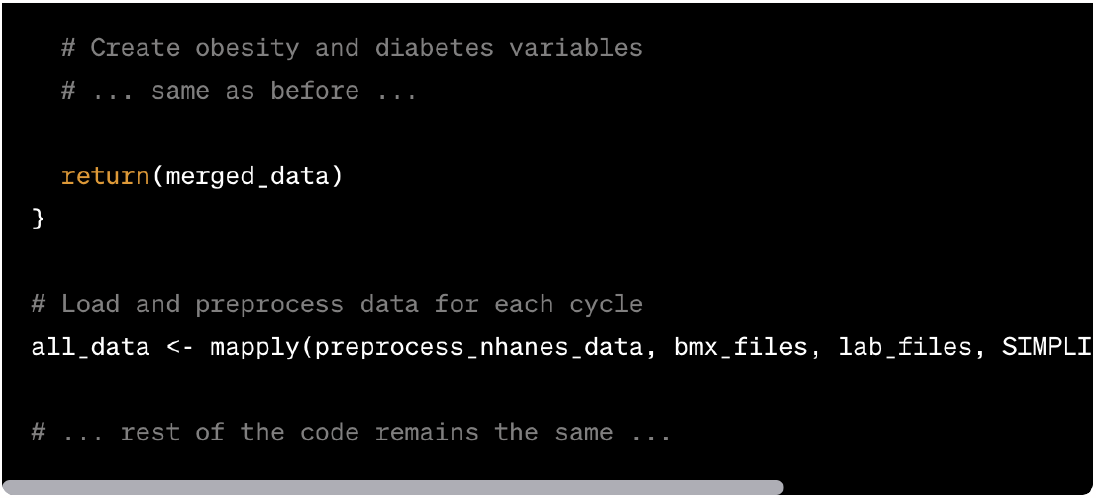

Remember to replace **“path/to/BMX_D0506.csv”**· and · **“path/to/LAB_D0506.csv”**· with the actual paths to your downloaded files. Also, add paths for the other cycles you’re including in your analysis.

This approach requires you to manually download and specify the file paths for each dataset, but it provides more control and flexibility in handling the NHANES data.

### You

Ok, looks like I had to download the diabetes data (DIQ), and I downloaded the body measures (BMX) as well. They are in a data folder in the same director as my R script. I have cycles 2005-2006 through 2017-2018. Labeled as XXX_{D-J} respectively. Now write me the data preparation code you mentioned earlier.

### NHANES-GPT

Great! With the Diabetes (DIQ) and Body Measures (BMX) datasets downloaded for the NHANES cycles from 2005-2006 to 2017-2018, we can proceed with the data preparation phase. I’ll provide you with the R code to load these datasets, merge them, and prepare them for analysis. Make sure your downloaded files are named consistently, like “DIQ_D.csv” for 2005-2006, “DIQ_E.csv” for 2007-2008, and so on.

Here’s the updated R code for data preparation:

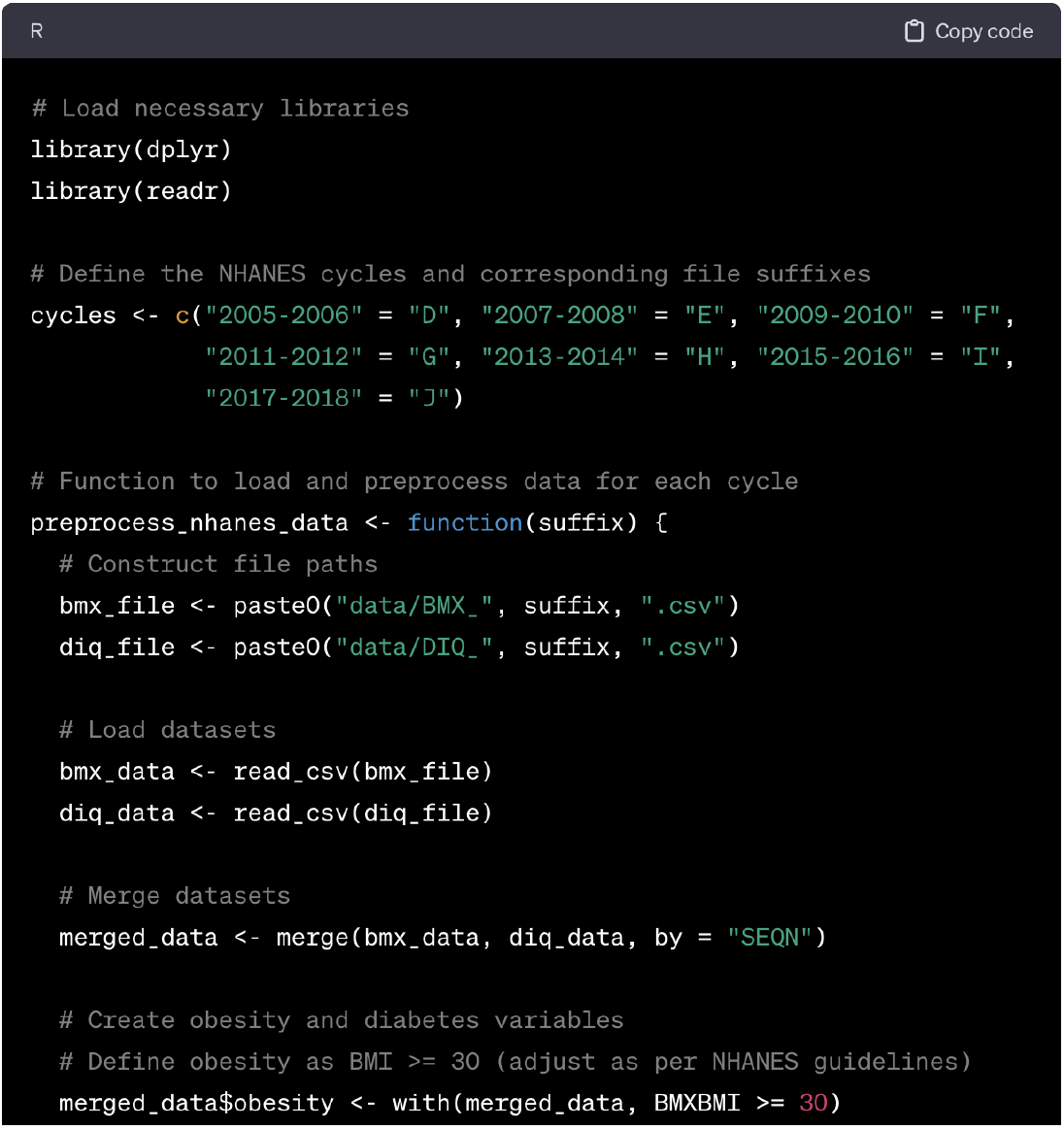

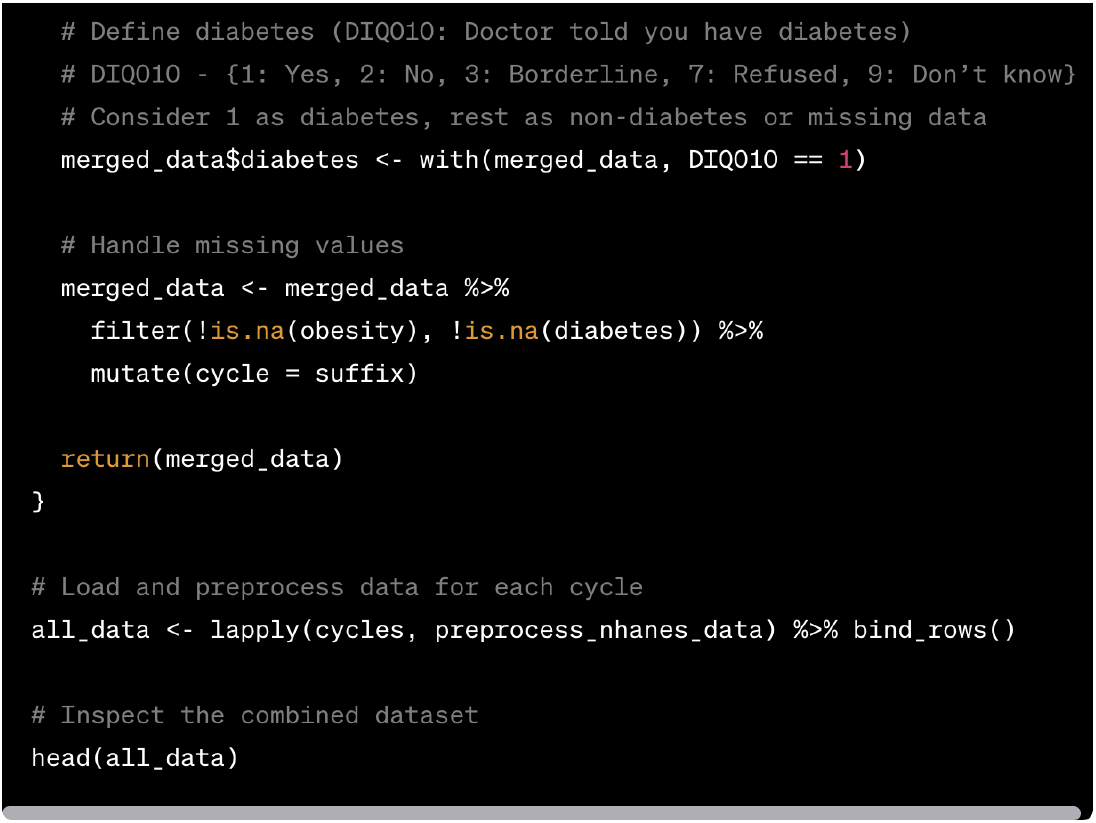

## Explanation

1. **Libraries:**We are using `‘dplyr’` for data manipulation and `‘readr’` for reading CSV files.
2. **Cycles and File Paths:** The `‘cycles’` vector maps each cycle to its corresponding uffix. The file paths are constructed based on these suffixes.
3. **Data Loading and Merging:** For each cycle, the corresponding BMX and DIQ files are loaded and merged on the unique identifier `‘sEQN’`
4. **Variable Creation:** We define obesity based on BMI (≥ 30) and diabetes based on the DIO010 variable (1 indicating diabetes).
5. **Missing Data:** We filter out rows where obesity or diabetes status is missing and add a new column to indicate the cycle.
6. **Data Inspection:** ‘`head(all_data)’` allows you to inspect the first few rows of the combined dataset.

Make sure to replace · **“**`data/BMX_`” and · **“**`data/DIQ_`” with the actual paths to your downloaded files, if they are different. Also, adjust the variable names and conditions based on the exact structure and coding of your NHANES datasets.

### You

Sorry, I forgot to mention these are all XPY files. How does that change things? The data is also in data/body _measures and data/diabetes, respectively.

### NHANES-GPT

Thanks for the update. XPY files are SAS transport files commonly used for NHANES data. To work with these files in R, you’ll need to use the `‘haven’` library, which can read SAS transport files. Let’s adjust the code to accommodate the XPY file format and the specified directory structure for the body measures and diabetes data.

First, make sure you have the `‘haven’` library installed. If not, you can install it using `‘install. packages (“haven”) ‘`.

Here is the modified R script:

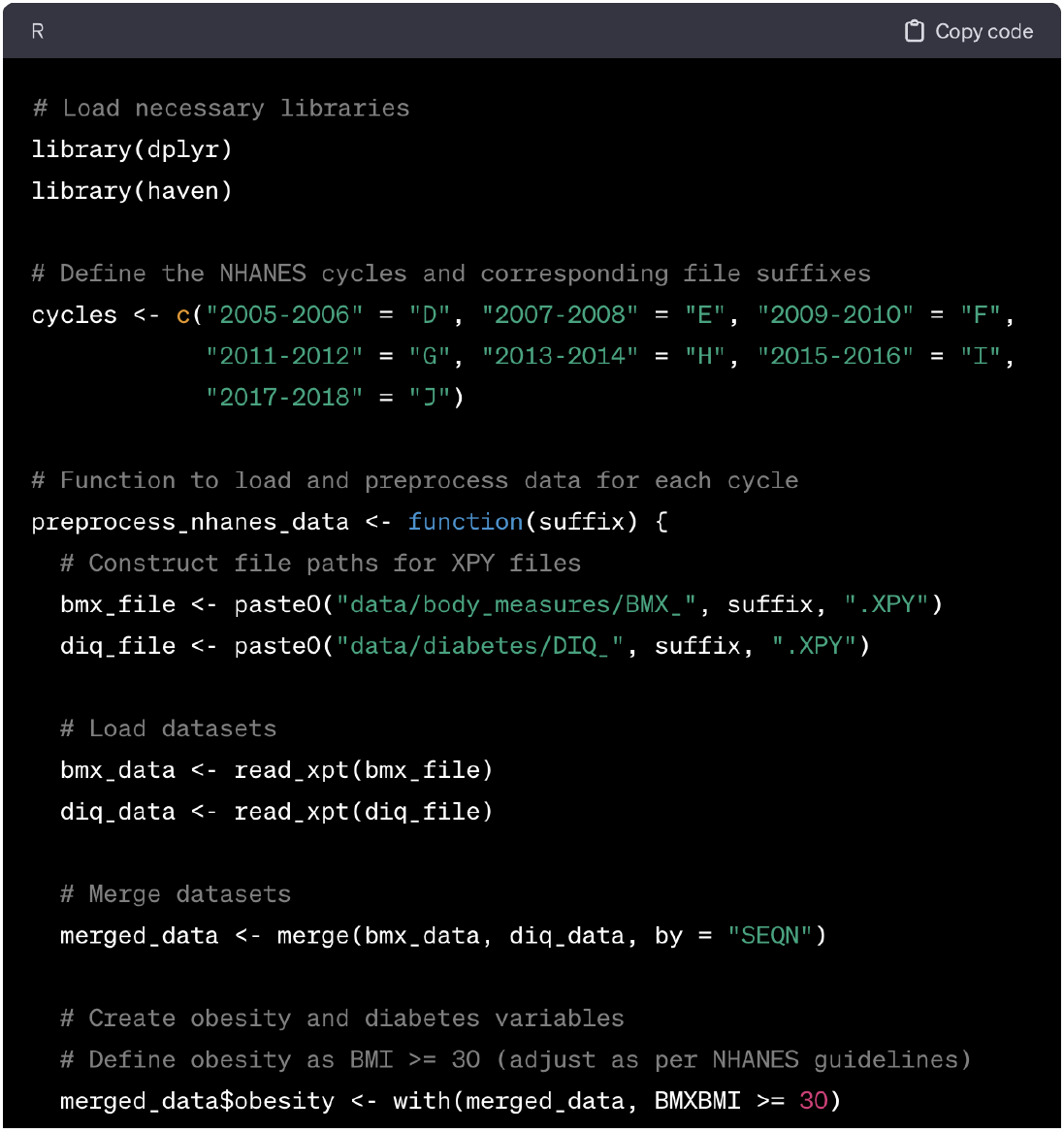

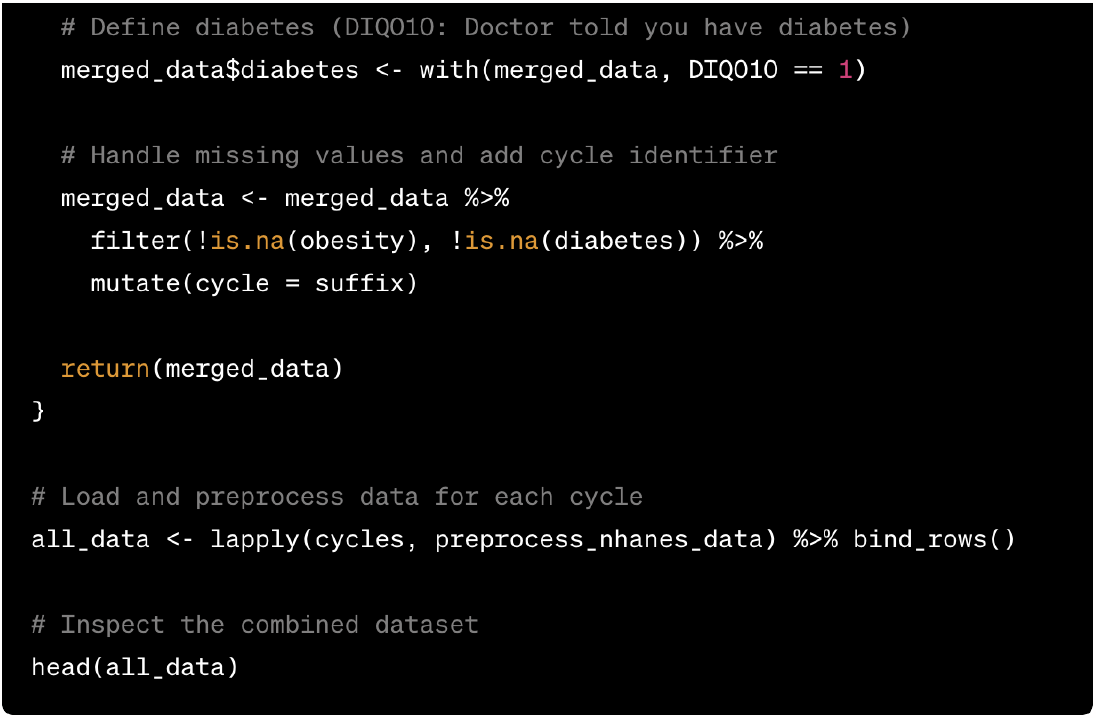

## Key Changes

1. **Library Change:** We now use `‘haven’` to read XPY files using `‘read_xpt()’`.
2. **File Paths:** The file paths are updated to point to the new locations and file formats `(“data/body_measures/BMX_”` and **“data/diabetes/DIQ_”**·).

Make sure the file paths and variable names match the actual files and contents. If there are any discrepancies in variable names or file formats, you’ll need to adjust the script accordingly.

### You

I got this error “Error: ‘data/body _measures/BMX_D.XPT’ does not exist in current working directory (‘/Users/titusmac’).” How do I change the working directory to the one my code is in?

### NHANES-GPT

To change the working directory in R to the directory where your R script is located, you can use the **‘setwd()’** function.However, if you want to set the working directory to the location of the script dynamically, especially if you move the script around, you can use the `‘here’` package, which is particularly useful for this purpose.

First, ensure you have the `‘here’` package installed. If not, install it using:

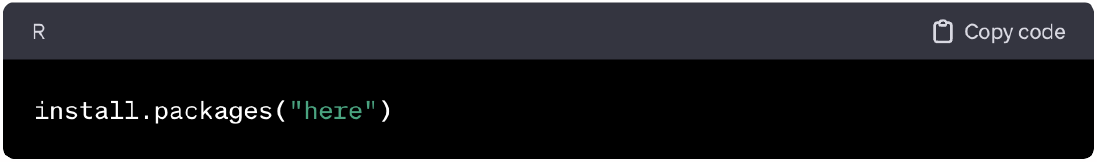

Then, use the `‘here’` function to dynamically set the working directory to the directory of your R script:

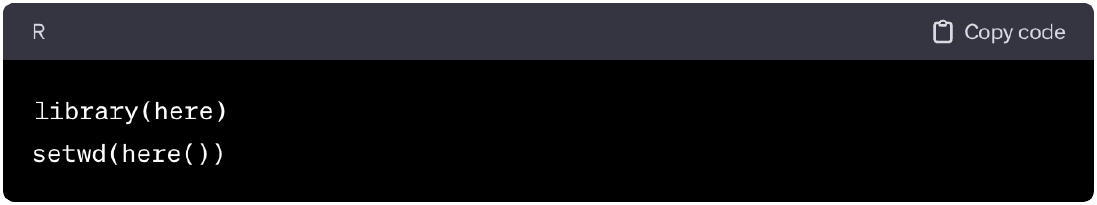

This will set the working directory to the location of your R script, making it easier to manage file paths. After setting the working directory, you can proceed with loading the XPY files as previously described. Remember, the paths to the XPY files should be relative to the location of your R script.

So your script should look something like this at the beginning:

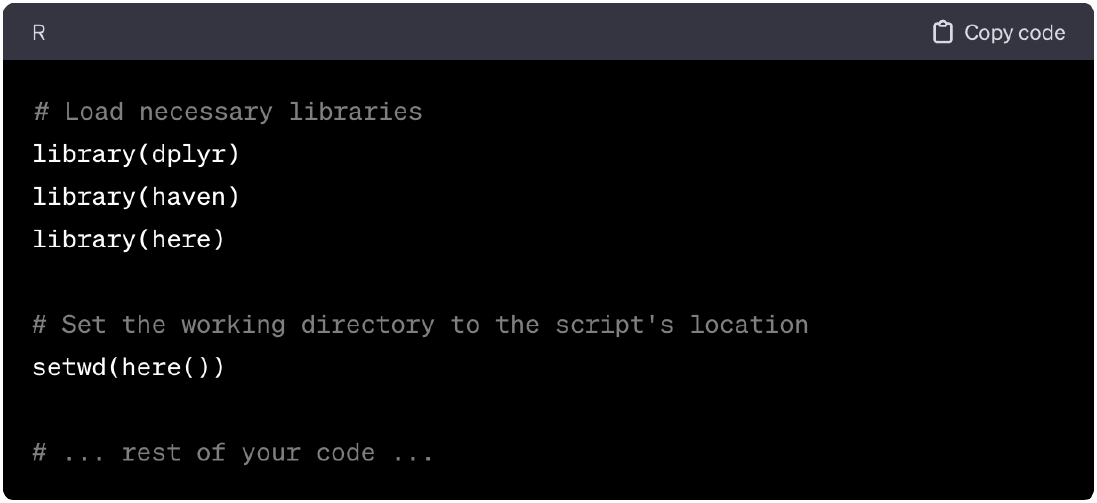

This should resolve the error you’re encountering, as long as the XPY files are in the correct locations relative to your script.

### You

Sorry, you also caught my type. They are in fact XPT files. But to the error above, here() still gives me the “/Users/titusmac” director, which is not the one I’m working in.

### NHANES-GPT

If the `‘here()’` function is not resolving to the correct directory, it might be due to the absence of a recognizable project structure, like an RStudio project or a directory with a ‘. `Rproj ‘` file. In such cases, `‘here () ‘` defaults to the current working directory as determined by R, which in your case seems to be `‘/Users/titusmac ‘`.

To set the working directory to the one containing your script, you can use an alternative approach:

### Manually Set the Working Directory

If you know the exact path of the directory, you can set it manually using `‘setwd() ‘`. For example:

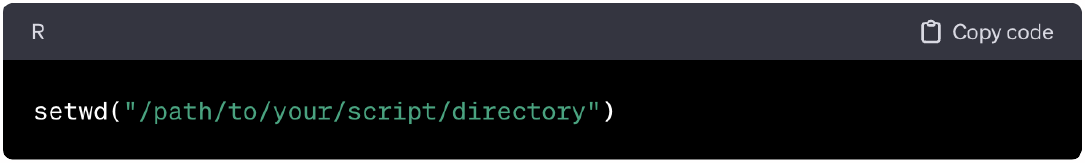

Replace `·”/path/to/your/script/directory”·` with the actual path where your script and data files are located.

**Using** `‘getwd() ‘` **and** `‘dirname() ‘` **with the Script File:**If you want to dynamically set the working directory to the location of the script, you can use the `‘dirname () ‘` function along with the script’s path. However, this requires you to know the script’s path. For example:

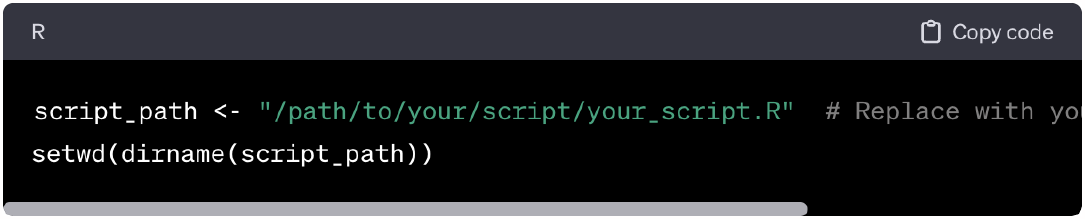

### RStudio Projects

If you are using RStudio, it’s a good practice to organize your work in RStudio projects. Each project sets the working directory to the project’s folder.

After setting the working directory, you can proceed with the rest of the script. The file paths for the XPT files should be relative to this directory.

Remember to replace the placeholders in the script with the actual paths relevant to your setup.

### You

I got through this step”# Inspect the combined dataset head(all_data)” and it seems like I have a lot of NAs.

### NHANES-GPT

If you’re encountering a large number of NAs (missing values) in your combined dataset, it suggests that there might be missing data in either the BMI (Body Mass Index) or diabetes variables, or possibly in other variables used in the merging process. Here are a few steps you can take to investigate and handle these missing values:

### Inspect Missing Data

First, understand where the missing values are. Are they concentrated in certain cycles, certain variables, or spread throughout?

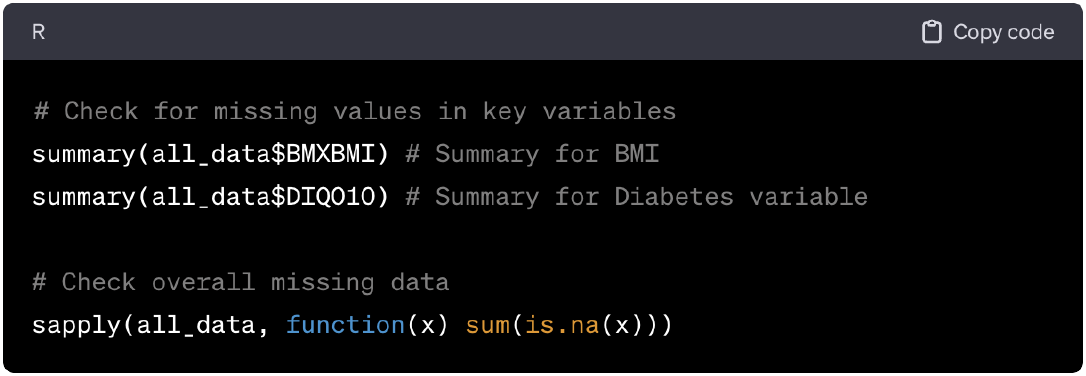

### Review Merging Process

Make sure that the merging of datasets by `‘SEQN’` is correct. Mismatches in `‘SEQN’` between datasets can lead to NAs.

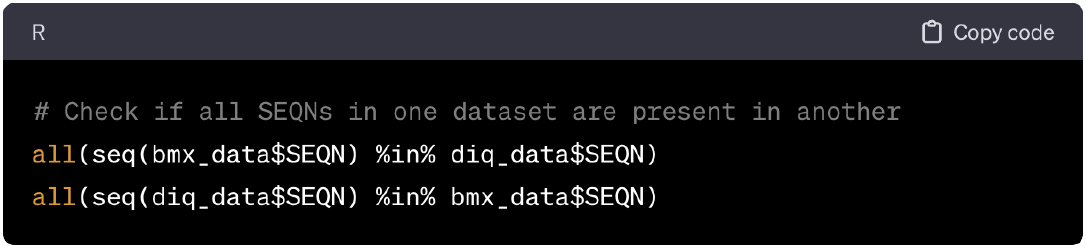

Handle Missing Data

### You have a few options to handle missing data

- **Exclude Rows with Missing Data:** If the number of rows with missing data is small, you might exclude them.

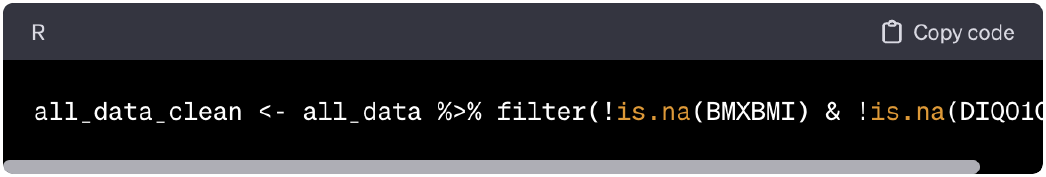

- **Imputation:** If the missing data is significant, consider imputing missing values. This is a more complex process and depends on the nature of your data and analysis.
- **Reconsider Variable Definitions:** Review how you are defining `‘obesity’` and **‘diabetes’**. Ensure that the thresholds and conditions are correctly applied.

### Further Analysis

If you’re still encountering issues, you may want to dive deeper into each dataset:

- Look at each dataset individually to understand the pattern of missing data.
- Check NHANES documentation for any known issues or peculiarities with the data.

Remember, handling missing data is a crucial step in data analysis and can significantly impact your results. The approach you choose should align with your research question and the nature of your data.

### You

Ok, I seem to have everything I need for those two variables. I just have a lot of NA in other variables, but I think we’re good to go. Now that I have the data pre-processing done, whats next?

### NHANES-GPT

Great! Now that you have successfully pre-processed the data, focusing primarily on the obesity and diabetes variables and managing the NAs in other variables, the next steps are:

### Incorporating NHANES Survey Weights and Complex Survey Design

NHANES uses a complex, multi-stage survey design. It’s important to incorporate this into your analysis to get nationally representative estimates.

### Statistical Analysis

This involves choosing appropriate statistical methods to analyze the trends in obesity and diabetes across different NHANES cycles.

### Interpreting Results

Understanding the output of your statistical analyses in the context of public health.

### Visualization

Creating visual representations of your findings can be highly beneficial for better understanding and communication of your results.

Let’s move on to these steps:

## 1. Incorporating NHANES Survey Weights and Complex Survey Design

NHANES provides sample weights (‘**WTMEC2YR**’ for medical examination weights, for example) that you need to use in your analysis to account for the complex survey design:

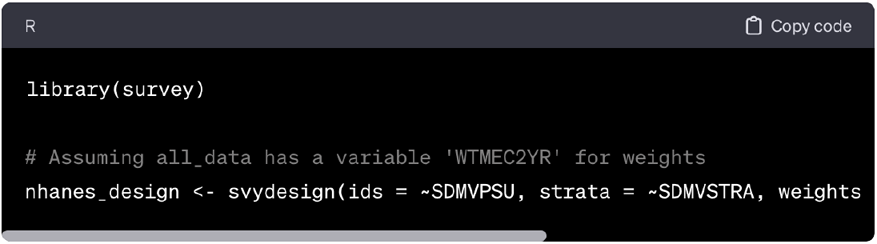

## 2. Statistical Analysis

You might want to perform logistic regression to assess the odds of obesity and diabetes over the cycles:

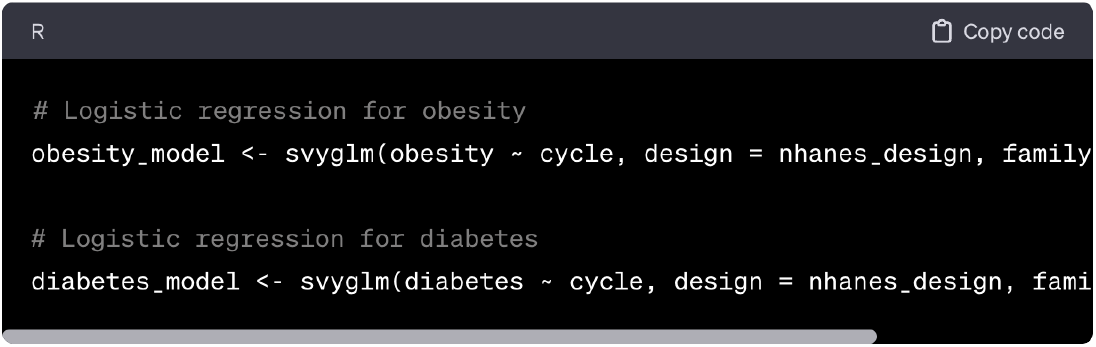

## 3. Interpreting Results

After running the models, you should look at the coefficients, p-values, and confidence intervals to understand the trends:

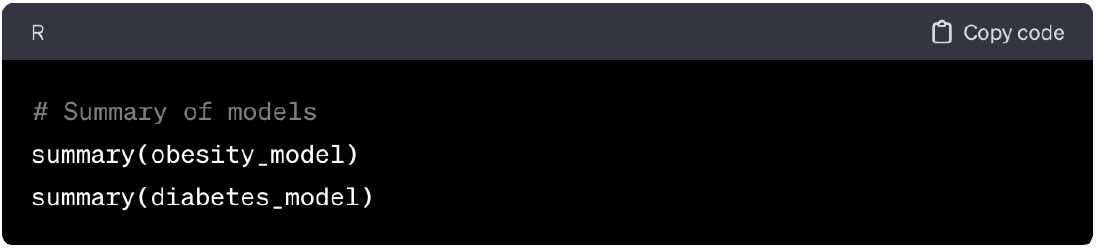

## 4. Visualization

Creating trend graphs over the cycles can be insightful:

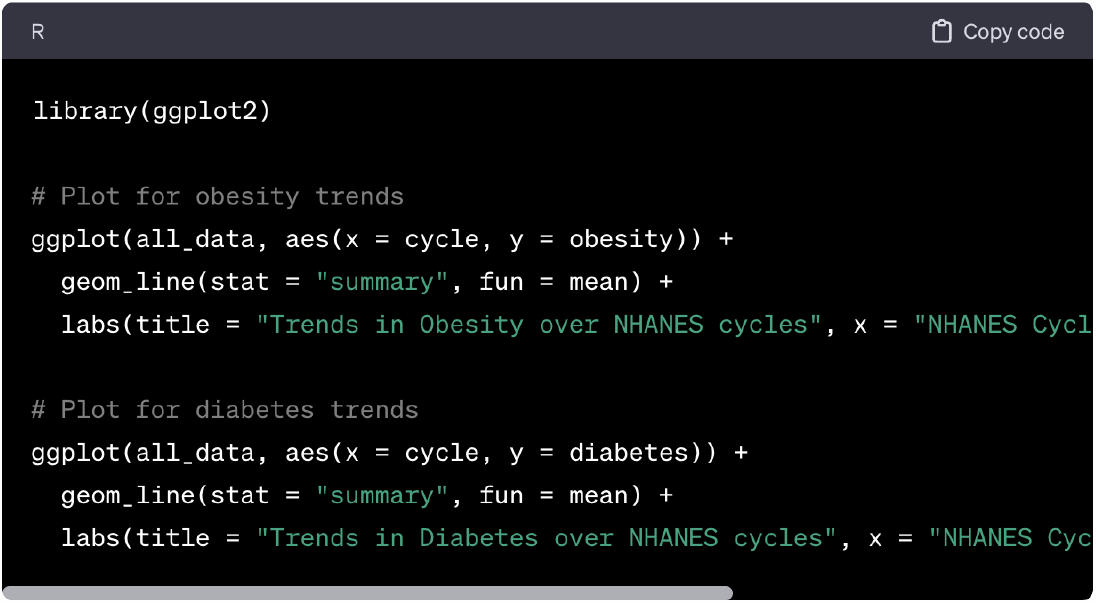

### Important Notes

- **Confounding Factors:**Consider adjusting your models for potential confounders like age, gender, race, etc.
- **Assumptions of Logistic Regression:** Make sure the assumptions for logistic regression are met.
- **Interpretation:**Carefully interpret the results, considering the limitations of the dataset and the analysis.

This should provide a comprehensive analysis of the trends in obesity and diabetes using the NHANES data. Remember, each step is crucial for a robust and valid analysis.

#### You

You didnt tell me to download the data with SDMVPSU or WTMEC2YR in it, so I got this error “Error in eval(predvars, data, env): object ‘SDMVPSU’ not found”

### NHANES-GPT

Apologies for the oversight. The variables **‘SDMVPSU**’ and **‘SDMVSTRA**’ are used for the primary sampling units and strata in the NHANES dataset, while **‘WTMEC2YR**’ is a sample weight variable. These are essential for analyzing NHANES data using the complex survey design.

You will need to download the Demographics Files for each of the NHANES cycles you’re working with, as these files contain the sample design variables (‘**SDMVPSU**’ and **‘SDMVSTRA**’) and the weights `(‘WTMEC2YR’` or similar).

Here are the steps to include these variables in your analysis:

### Download Demographic Data

Go to the NHANES website and download the Demographic files (e.g., named as ‘**DEMO _XXXX. XPT**’) for each cycle you are analyzing.

### Merge Demographic Data with Your Existing Dataset

You’ll need to merge the demographic data with your previously combined obesity and diabetes data. The merging should be based on the ‘**SEQN**’ variable, which is the unique identifier for each respondent.

### Update the Analysis Code

After merging, you’ll have access to the necessary variables for the complex survey design analysis.

Here’s a revised version of the part of your script that merges the demographic data:

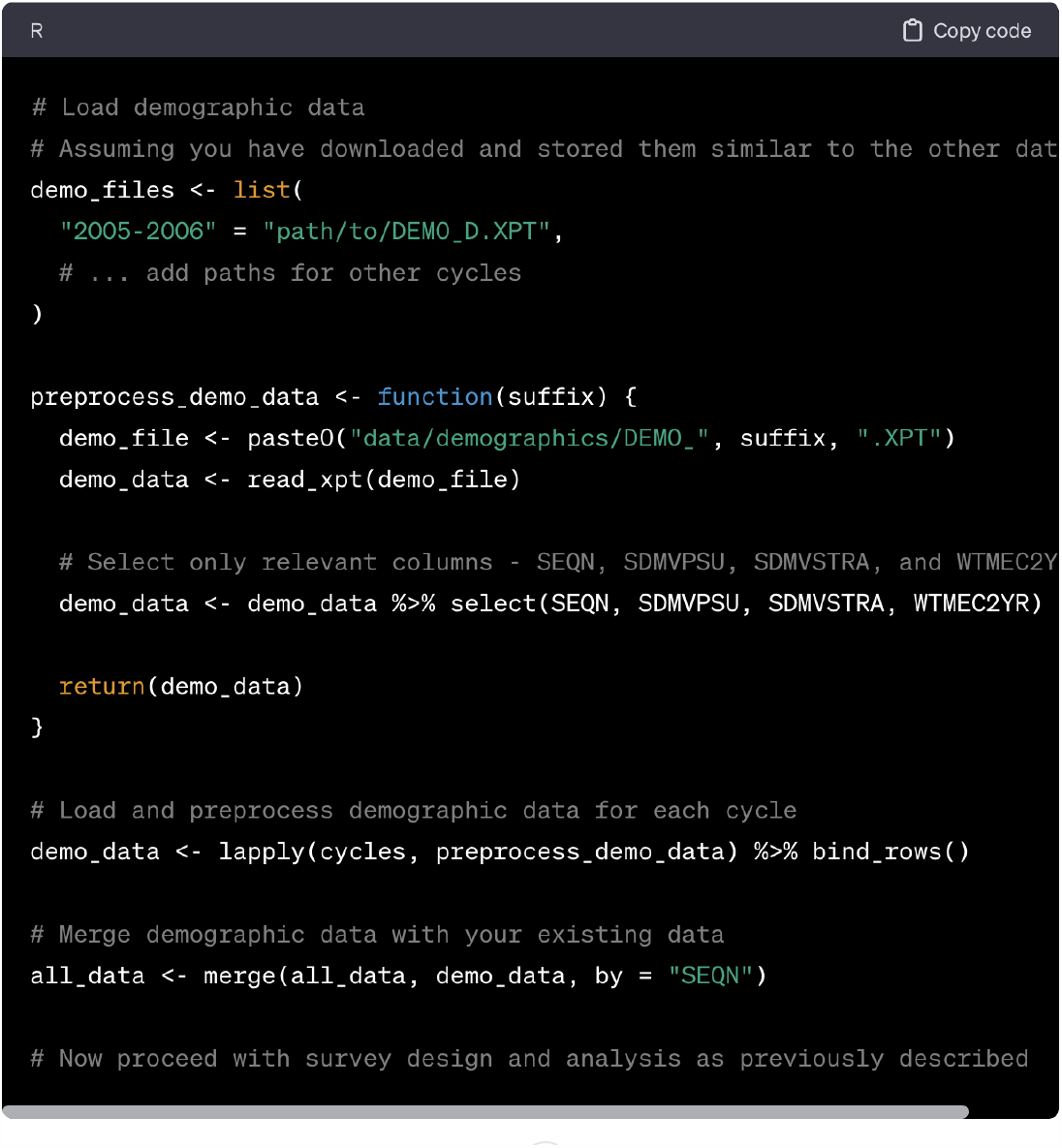

Remember to replace · **“path/to/DEMO_D.XPT”**· with the actual path where you’ve stored the demographic data files. This addition should resolve the error and allow you to incorporate the complex survey design into your analysis.

#### You

Its the exact same file structure as the last two, cant I just update that code?

### NHANES-GPT

Certainly, you can update the existing data processing code to include the demographic data. Since the file structure is the same as the body measures and diabetes data, you’ll just need to add a few lines to read and merge the demographic data.

Here’s the updated code:

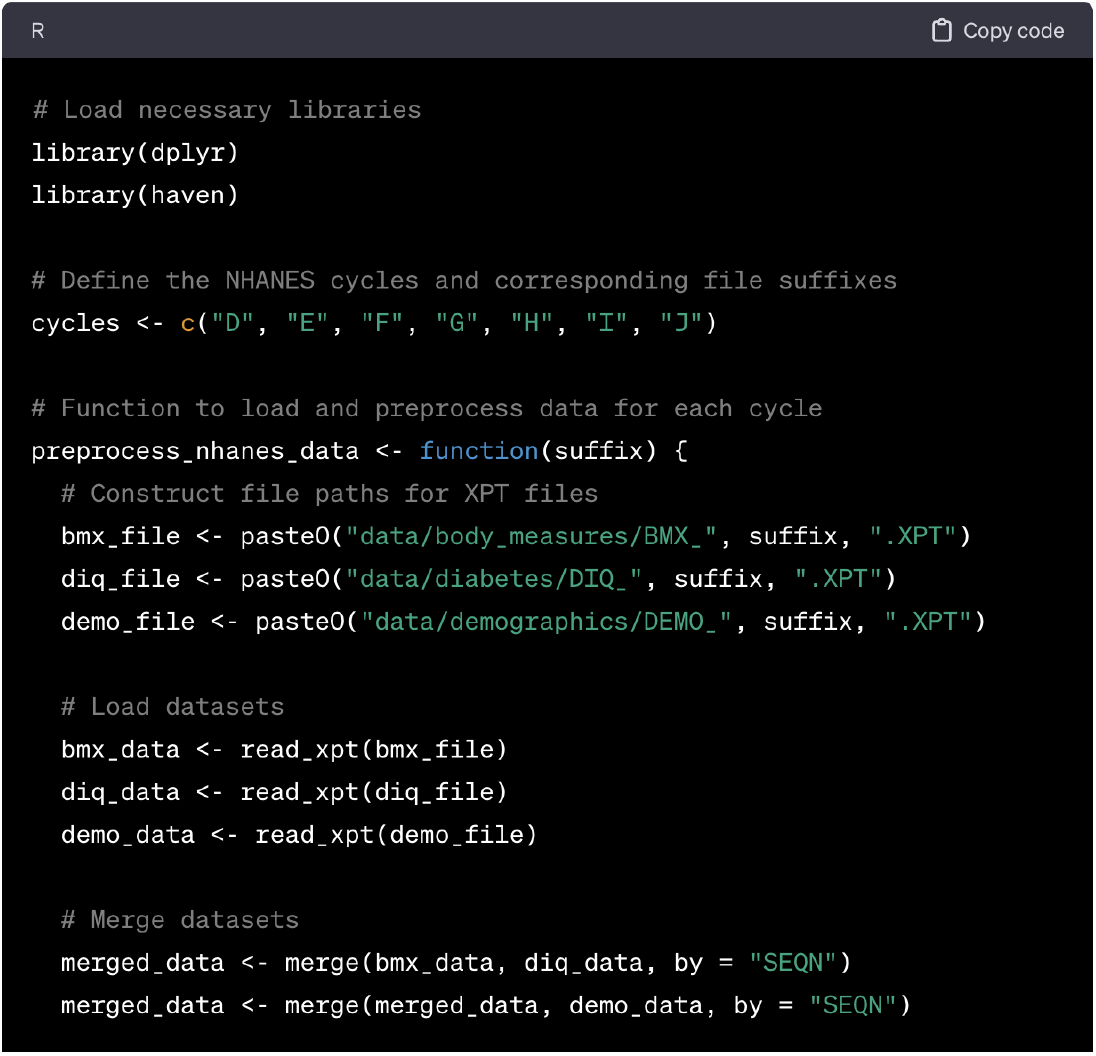

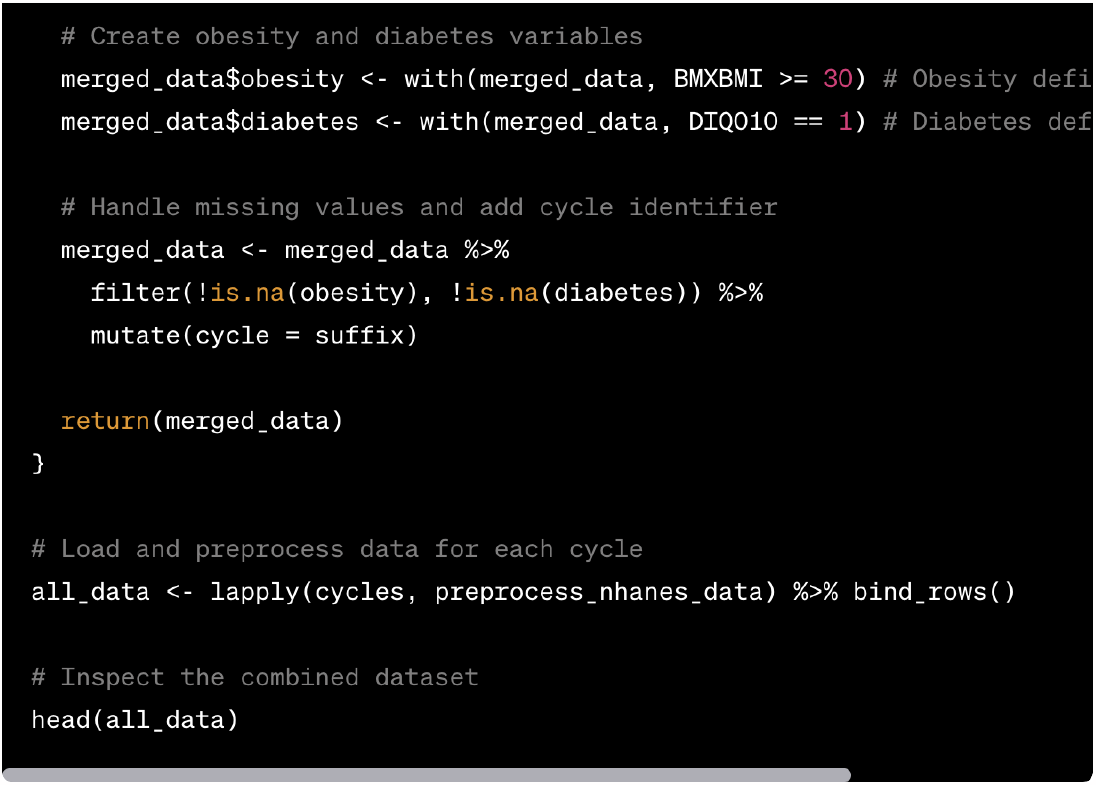

This updated code will read and merge the demographic data along with the body measures and diabetes data. Make sure the variables `‘SDMVPSU’, ‘SDMVSTRA’`, and `‘WTMEC2YR’` (or their equivalents, depending on the NHANES cycle) are present in the demographic datasets.

Once you have the combined dataset with all the necessary variables, you can proceed with the survey weighting and complex survey design analysis as previously described.

#### You

Ok, survey design worked with the DEMO data. Whats next?

### NHANES-GPT

Great to hear that the survey design integration worked with the demographic data. The next steps are statistical analysis, interpretation of the results, and data visualization. Let’s proceed with these:

## 1. Statistical Analysis

Given the nature of the data, logistic regression is a suitable choice for analyzing the trends in obesity and diabetes. Since NHANES uses a complex survey design, you’ll use functions from the **`‘survey’`** package to account for this in the analysis.

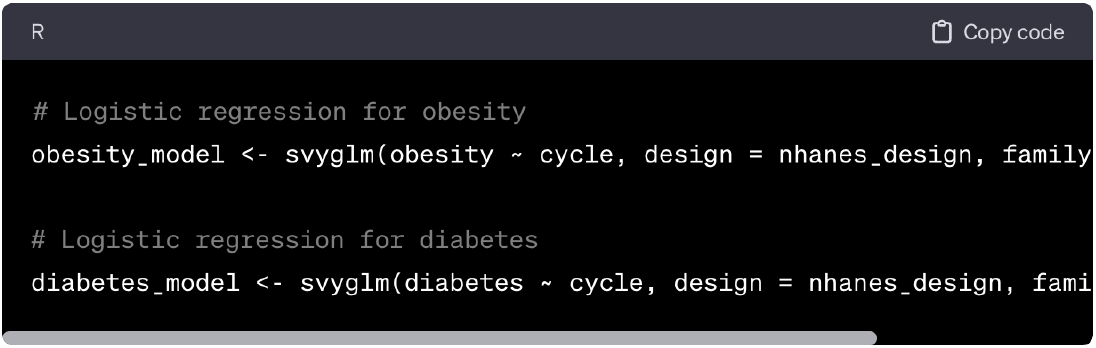

## 2. Interpreting Results

After running the models, examine the summary of each model to understand the relationship between the NHANES cycle and the prevalence of obesity and diabetes. Look for trends over time and the significance of these trends.

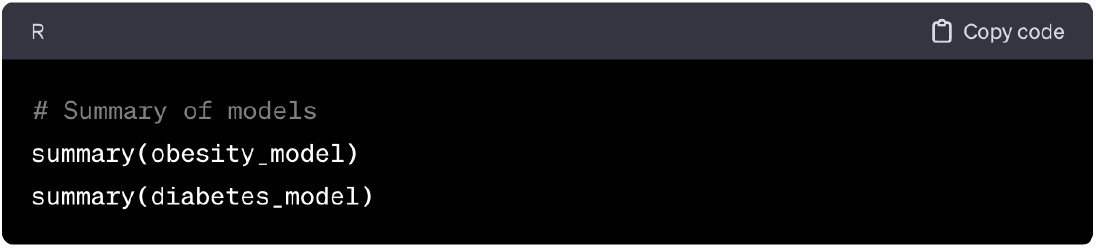

## 3. Visualization

Create visual representations of your findings. Graphs can help in illustrating the trends across different cycles.

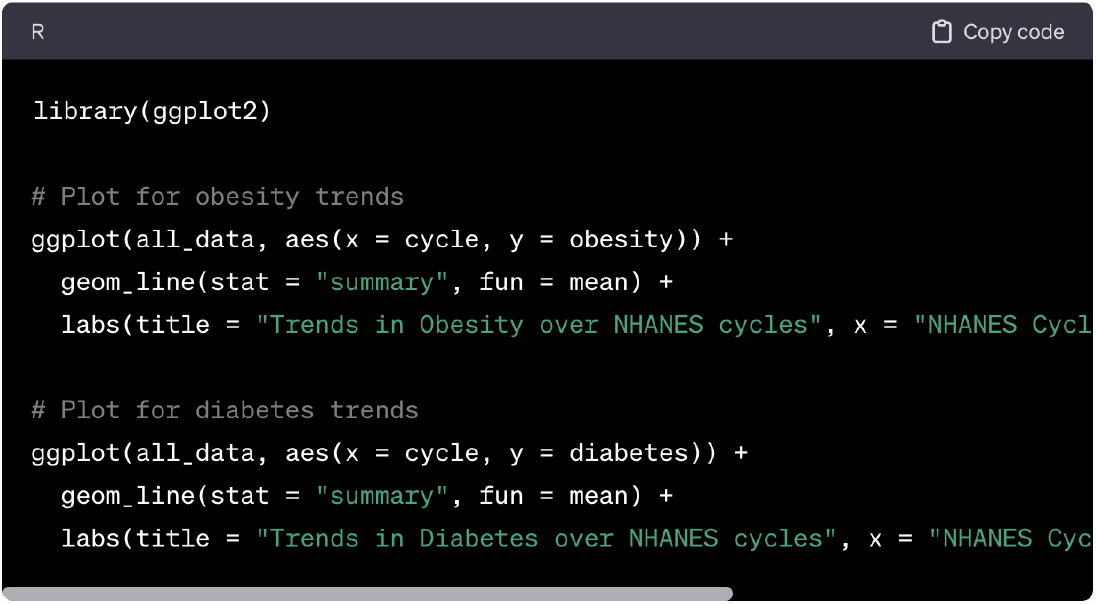

## Additional Considerations

- **Confounding Factors:**Consider adjusting your models for potential confounders like age, gender, race, etc.
- **Check Assumptions:** Ensure that the assumptions for logistic regression are met.
- **Interpretation:**Interpret the results carefully, considering the limitations of the dataset and the analysis.
- **Further Analysis:** Depending on the results, you might want to delve deeper into specific subgroups or related variables.

These steps should provide a comprehensive analysis of the obesity and diabetes trends in the NHANES data. Remember, the interpretation of the results should be done in the context of the broader public health landscape and existing literature.

### You

With the two models, I got this warning. What does it mean? “Warning message: In eval(family$initialize): non-integer #successes in a binomial glm!”

### NHANES-GPT

The warning “non-integer #successes in a binomial glm!” typically occurs in logistic regression (generalized linear models with binomial family) when the response variable is not an integer. In the context of logistic regression, the response variable is expected to be a count of successes and failures (typically Os and1s for binary outcomes).

Given that your analysis is based on NHANES data with ‘obesity’ and ‘diabetes’ as response variables, this warning suggests that these variables might not be strictly coded as Os and 1s. Let’s troubleshoot this:

### Check Data Types

Ensure that ‘obesity’ and ‘diabetes’ are binary (0 or 1) and are stored as integers. If they are coded differently, recode them accordingly.

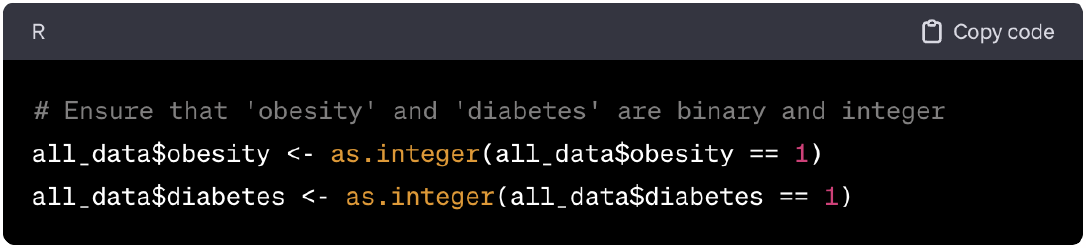

### Fractional Values

If ‘obesity’ and ‘diabetes’ are calculated as proportions or have fractional values, this can cause the warning. They should be individual-level binary indicators.

### Aggregated Data

If you’re working with aggregated data (like means or proportions), logistic regression might not be appropriate. Logistic regression is designed for individual-level binary outcome data.

After making these checks and adjustments, rerun the logistic regression models. If the issue was with the coding of the response variables, these modifications should resolve the warning. If the warning persists, it might be necessary to further investigate the nature of the data you’re working with and ensure it’s appropriate for logistic regression.

### You

When I run the ggplot code, I get this error”‘geom_line()’: Each group consists of only one observation.

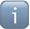Do you need to adjust the group aesthetic?”

### NHANES-GPT

The error you’re encountering with ‘`geom_line()’` in ggplot2 usually occurs when ggplot is unable to find multiple observations per group to connect with lines. This is often due to a misunderstanding of how ggplot interprets the data, especially regarding how it defines groups.

In your case, the issue likely stems from the way the data is being passed to ggplot for plotting trends over NHANES cycles. Since your objective is to plot trends over time, you need to ensure that the data is grouped and summarized correctly before plotting.

Here’s an adjusted approach:

### Create a Summary Dataset

Before plotting, create a summary dataset where you calculate the mean (or proportion) of obesity and diabetes for each cycle.

### Plot Using the Summary Dataset

Use this summarized dataset to plot the trends.

Here’s how you can modify the code:

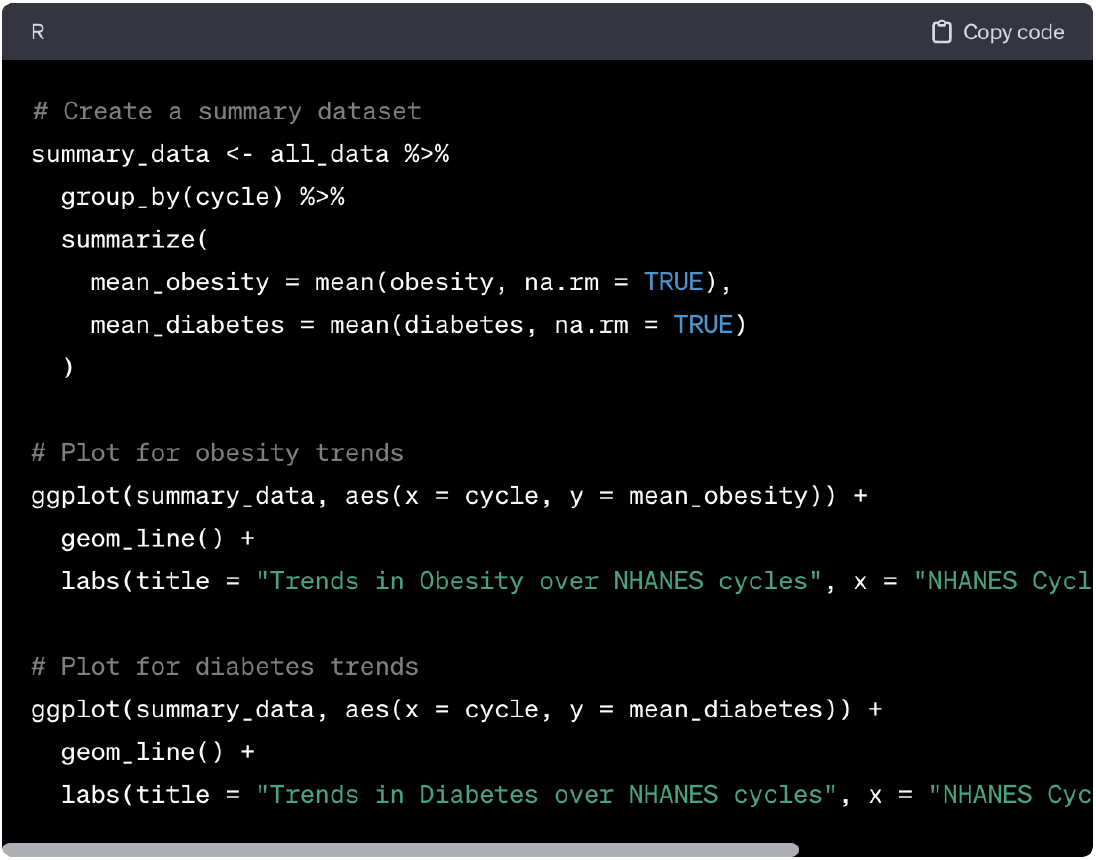

In this revised code, ‘`summary _data’` is a new dataframe where the mean prevalence of obesity and diabetes is calculated for each NHANES cycle. This data is then used to plot the trends, with **‘geom_line()**’ connecting the mean values across cycles.

This approach should resolve the error and allow you to visualize the trends in obesity and diabetes across NHANES cycles effectively.

#### You

Hmm, its still giving me the same error.

### NHANES-GPT

I see, let’s troubleshoot this further. The error message suggests an issue with the grouping of data for the line plot in ggplot2.Since ‘`geom_line()’` requires a grouping variable to connect points, we need to ensure that ggplot understands how to group your data.

In your case, since you’re trying to plot trends over time (cycles), the `‘cycle’` variable should be the grouping variable. However, ggplot might not be automatically recognizing `‘cycle’` as a discrete variable suitable for grouping. Let’s explicitly set the `‘group’` aesthetic in the ggplot call to resolve this issue:

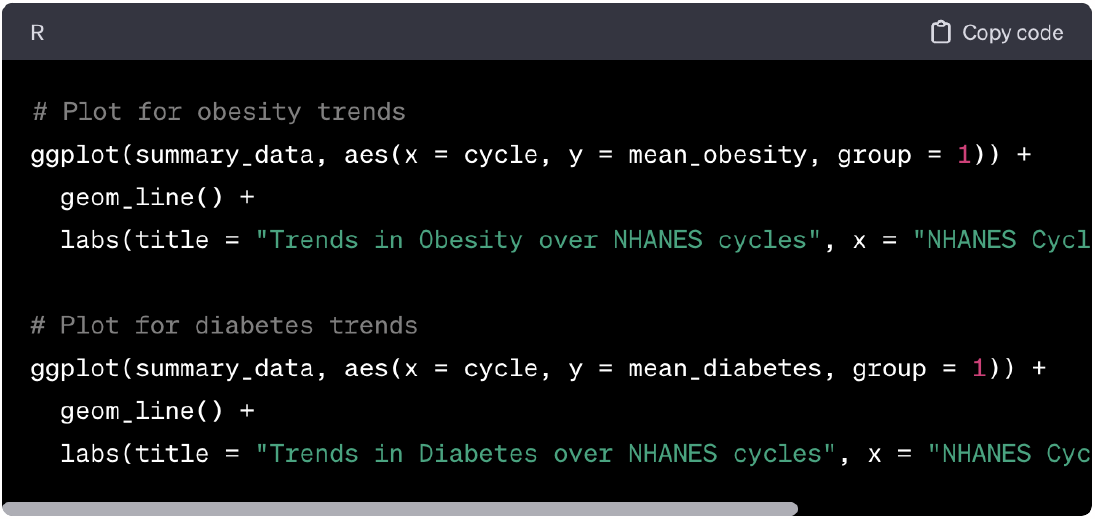

In this revised code, I’ve added `‘group` = **1**’ to the `‘aes()’` function in ggplot. This explicitly tells ggplot to consider all data as part of a single group for the purposes of plotting lines. This should resolve the issue and allow your line plots to render correctly.

If the error still persists, there might be a more fundamental issue with the structure of your `‘summary _data’` dataset or how `‘cycle’` is formatted. In that case, you would need to inspect `‘summary _data’` and ensure that `‘cycle’` is correctly formatted and that there are multiple observations for each cycle.

#### You

That worked!. But can we update this to be a more attractive plot using tidyverse best practices?

### NHANES-GPT

Absolutely! Creating a more visually appealing and informative plot using tidyverse best practices is a great idea. Let’s enhance the readability and aesthetics of your plots. Here’s how you can improve the plots for obesity and diabetes trends:

#### Use Readable Labels

Improve axis labels and titles for better understanding.

#### Add Points for Each Observation

This helps to visualize individual data points.

#### Use Themes

Apply a theme for a cleaner look.

#### Color and Size Adjustments

Make lines and points more visually appealing.

#### Facet Wrap (Optional)

If you want to display both obesity and diabetes trends in a single plot, you can use `‘facet_wrap’`.

Here’s an updated version of your ggplot code:

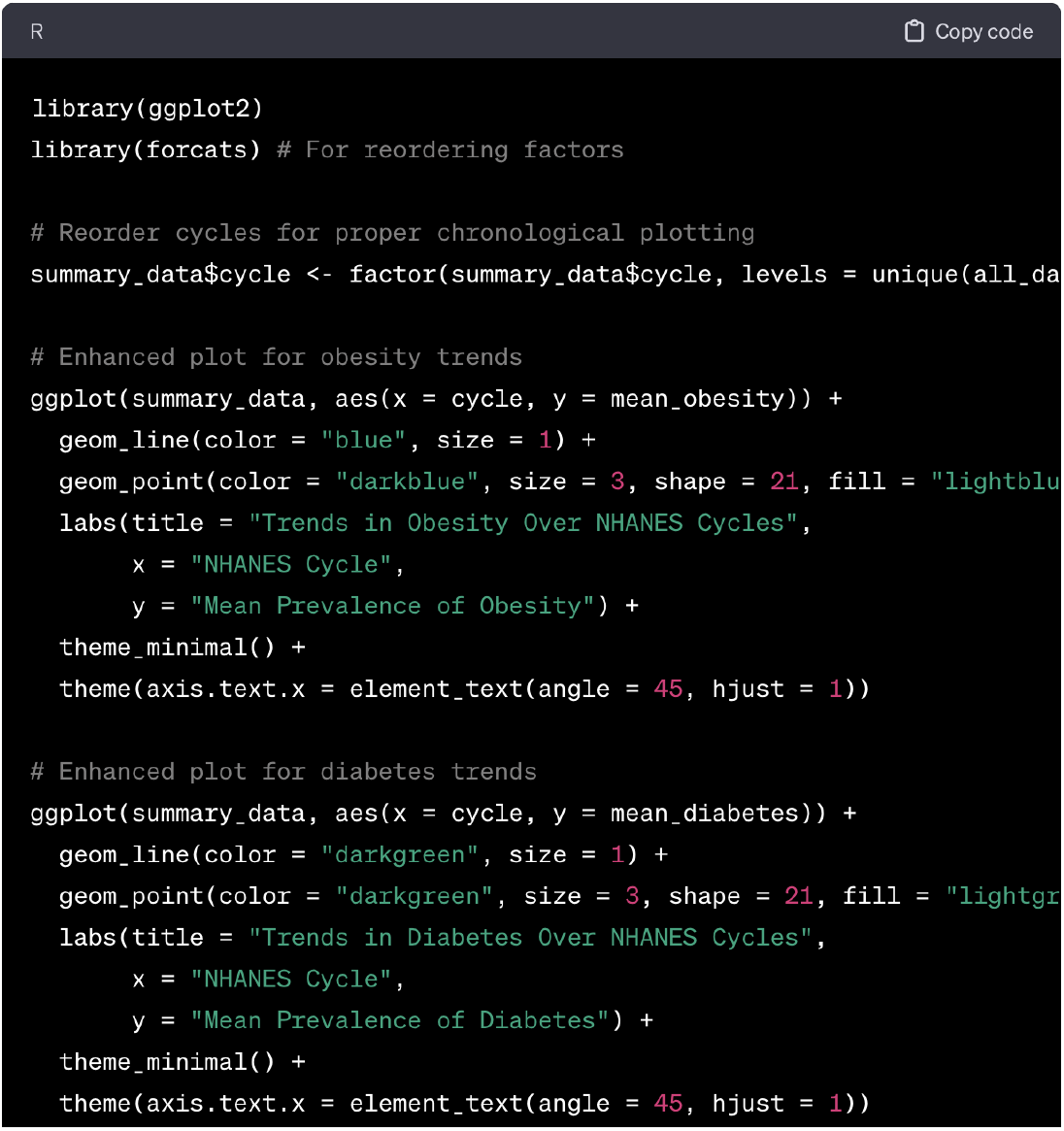

These enhancements should make your plots more attractive and easier to interpret. The `‘theme_minimal()’` provides a clean and modern look, and the adjusted axis labels and titles clarify the information being presented. The addition of points helps to highlight individual cycle data, and the color choices make the plots more visually appealing. Adjust colors and themes as per your preference.

#### You

These arent much better looking and I think its because I am getting the same group error. Can you explicitly cast those again in this code?

### NHANES-GPT

Certainly! Let’s reintroduce the `‘group’` aesthetic and further enhance the visual appeal of your plots. Ensuring that the `‘group’` aesthetic is correctly specified will address the grouping issue. Additionally, we can make a few more adjustments to improve the overall look of the plots.

Here’s the updated ggplot code with the `‘group’` aesthetic and additional styling enhancements:

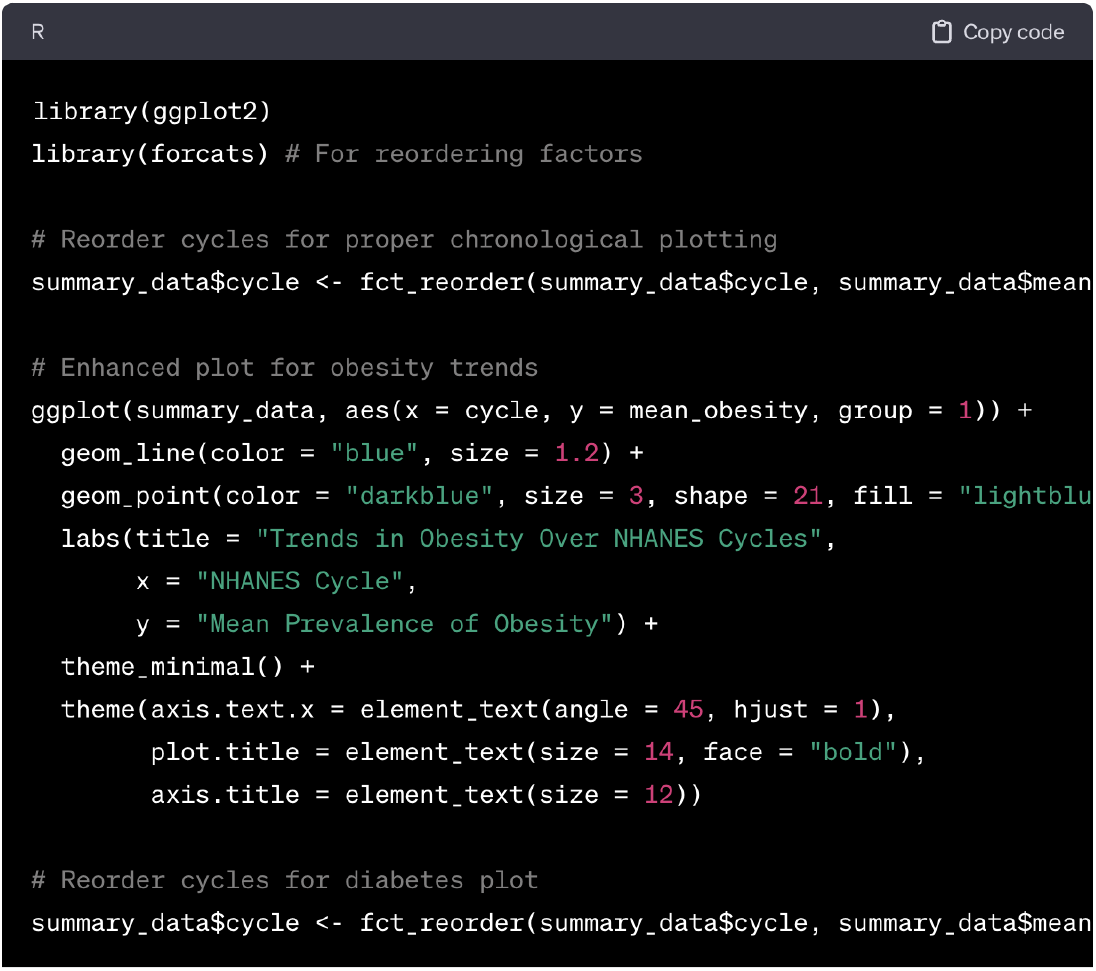

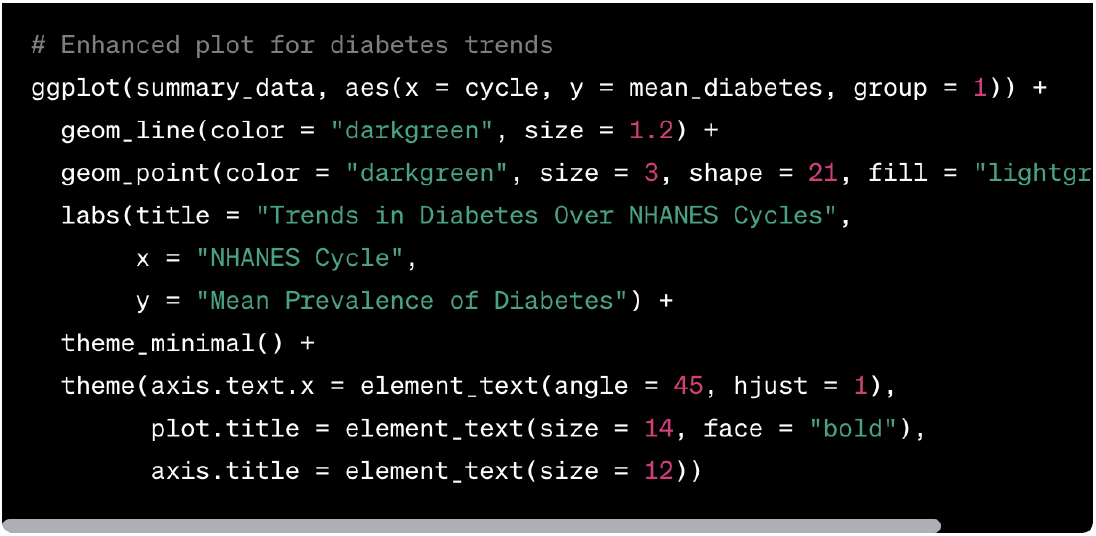

In this version:

- The `‘group` **=** **1**’ aesthetic is explicitly set in both `‘geom_line()’` and ‘**geom_point()’**.
- The `‘theme_minimal()’` is used for a clean look.
- Axis labels are rotated for better readability.
- The title and axis titles are emphasized with bold and larger font sizes.
- The `‘fct_reorder’` function is used to ensure the cycles are plotted in the correct order.

These changes should create more visually appealing and informative plots, and explicitly setting the `‘group’` aesthetic should resolve the grouping issue.

#### You

Lets relable the x-axis with survey years (e.g. 2005-2006) so its more understandable.

### NHANES-GPT

Absolutely, relabeling the x-axis with the full survey years (e.g., “2005-2006”) will make the plots more understandable. To do this, we will modify the `‘cycle’` variable in the ‘**summary _data’** dataframe to include the full survey years. Here’s how you can update the code:

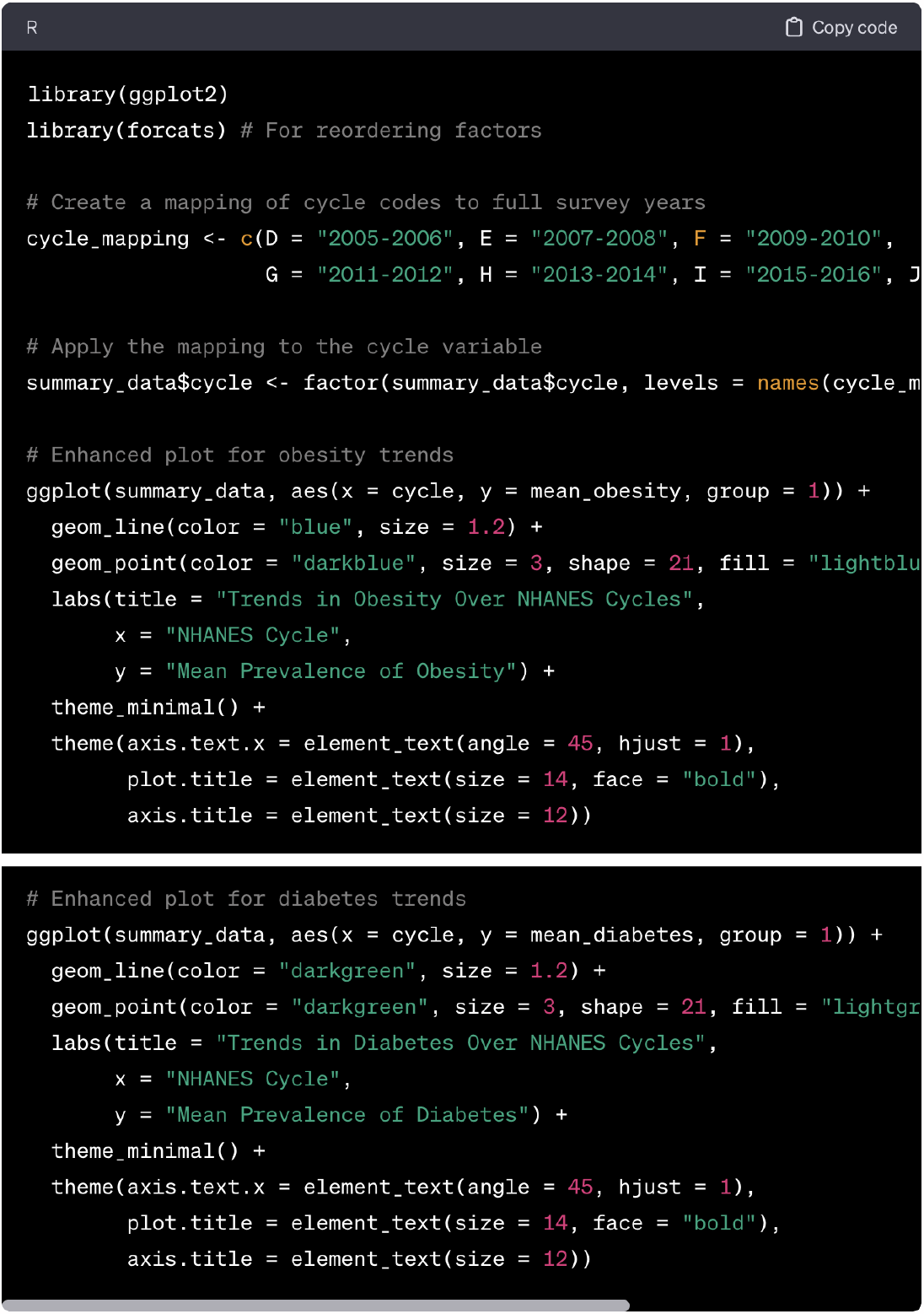

In this updated code:

- We define **‘cycle_mapping**’ to map the cycle codes (e.g., “D”, “E”, “F”) to the full survey years (e.g., “2005-2006”, “2007-2008”).
- We then apply this mapping to the `‘cycle’` variable in the `‘summary _data’` dataframe, converting it to a factor with appropriate labels.
- The plots will now display the full survey years on the x-axis, enhancing readability and understanding.

This should make the plots more user-friendly and informative for anyone interpreting the trends in obesity and diabetes over NHANES cycles.

#### You

Ok, last plotting request. I want these plots saved to a PNG with a DPI = 300 so they are publication ready.

### NHANES-GPT

Certainly! Saving your plots to a high-quality PNG file with a DPI of 300 is a great practice for ensuring they are ready for publication. You can use the **‘ggsave()**’ function from the ggplot2 package to do this. Here’s how you can save your plots:

First, you’ll need to store each plot in a variable, and then use **‘ggsave()**’ to save them.

Let’s modify the code accordingly:

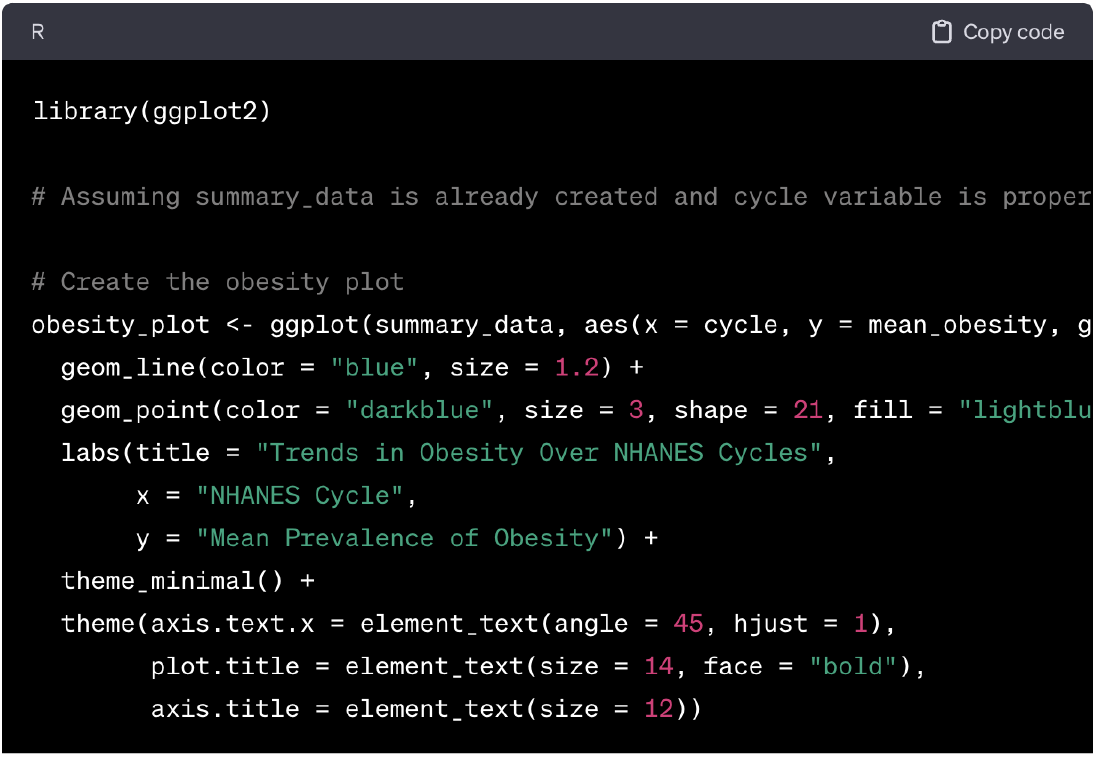

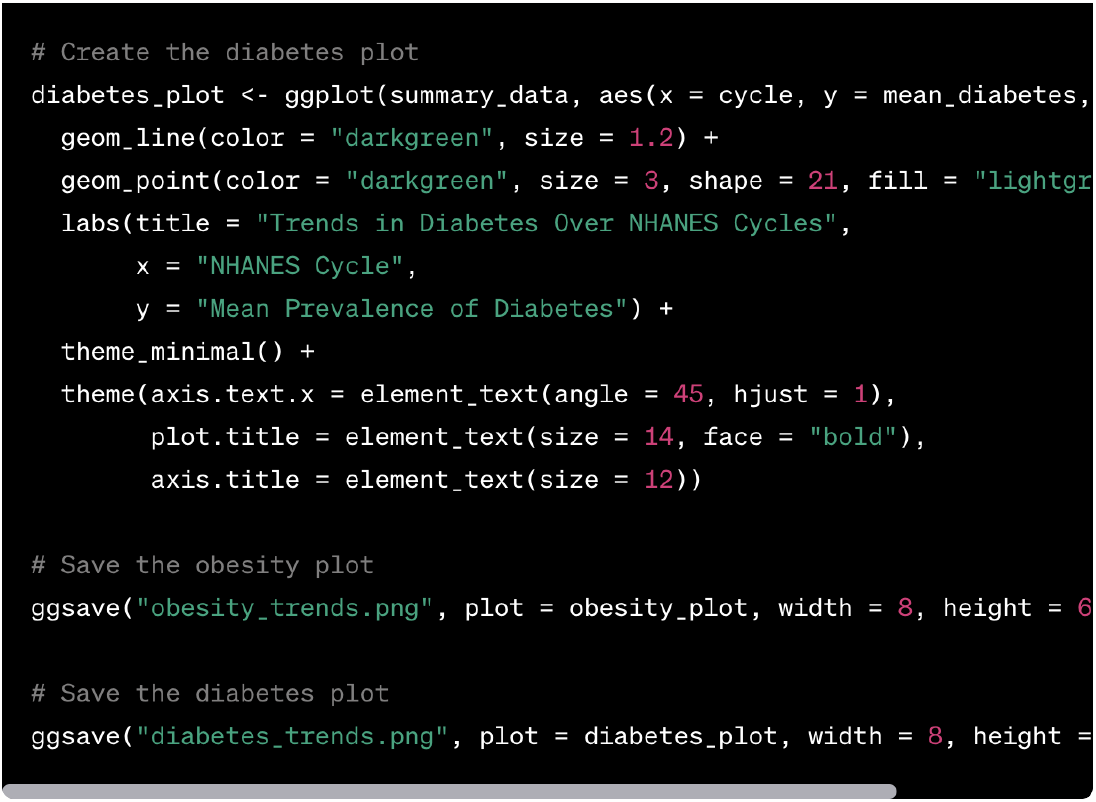

This code will create two PNG files: `‘obesity_ trends. png’` and `‘diabetes_ trends. png’`. They will be saved in your current working directory. The `‘width’` and `‘height’` parameters in `‘ggsave ()’` set the size of the saved image, and `‘dpi = 300’` ensures a high-resolution output suitable for publication.

Remember to check your current working directory to locate these files, or you can provide a full path to `‘ggsave()’` to specify a different save location.

#### You

Great, I think we’re wrapped up with this part, but I thought I would need to control for confounding variables in this process and we didn’t do any of that. What are the variables I should make sure to control for and why, and what survey data are they in and what are the NHANES-specific variable names?

### NHANES-GPT

Controlling for confounding variables is indeed an important aspect of epidemiological and public health research. Confounders are variables that are associated with both the exposure (in your case, the NHANES cycles) and the outcome (obesity and diabetes), and can bias the observed association between them. Including relevant confounders in your statistical model helps to provide a more accurate estimate of the relationship you are investigating.

For the analysis of obesity and diabetes trends using NHANES data, common confounders to consider include:

#### Age

Age is a critical factor in both obesity and diabetes prevalence.

#### Gender

There can be gender differences in obesity and diabetes rates.

#### Race/Ethnicity

Different racial and ethnic groups may have different rates of obesity and diabetes, often due to a combination of genetic, environmental, and socio economic factors.

#### Socioeconomic Status (SES)

Indicators of SES (like education, income) are often linked to health outcomes.

#### Smoking Status

Smoking has been associated with metabolism and appetite, and thus can be a confounder.

For these confounders, you can find the relevant variables in the NHANES Demographic (DEMO) and Questionnaire (e.g., Smoking - SMQ) data files. Here are the NHANES-specific variable names:

- **Age:** `‘RIDAGEYR’` (Age in years at screening) in the DEMO file.
- **Gender:** `‘RIAGENDR’` (Gender) in the DEMO file.
- **Race/Ethnicity:** `‘RIDRETH3’` (Race/Hispanic origin w/ **NH** white/Black/Asian) in the DEMO file.
- **Education Level** (as a proxy for SES): `‘DMDEDUC2’` (Education level - adults 20+) in the DEMO file.
- **Income Level** (another proxy for SES): `‘INDHHIN2’` (Annual household income) in the DEMO file.
- **Smoking Status:** `‘SMQ020’` (Smoked at least 100 cigarettes in life) in the SMQ file.

To control for these variables in your analysis, you’ll need to:

1. **Merge the Relevant Data:** Ensure these variables are included in your dataset during the data merging step.
2. **Modify Your Statistical Models:** Include these variables as covariates in your logistic regression models.

For example, the logistic regression model for obesity would be modified as follows:

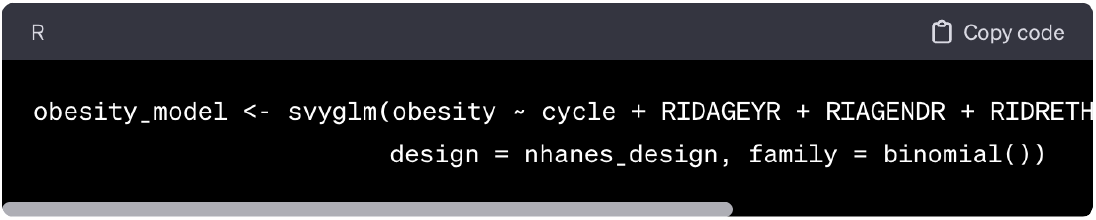

Remember, the inclusion of these variables as confounders should be guided by your research question and understanding of the subject matter. Additionally, you might need to recode or categorize some of these variables (like age or income) for appropriate inclusion in the models.

#### You

This is the model summary of the obesity model without controlling for confounding.

“Call:

svyglm(formula = obesity ∼ cycle, design= nhanes_design, family= binomial())

Survey design:

svydesign(ids = ∼SDMVPSU, strata= ∼SDMVSTRA, weights= ∼WTMEC2YR, data= all_data, nest= TRUE)

Coefficients:

Estimate Std. Error t value Pr(>ltl)

(Intercept) -0.77948 0.06450-12.085 < 2e-16 ***

cycleE--0.02715-0.08181 -0.332 0.740670

cycleF-0.06827-0.07541 0.905 0.367422

cycleG-0.04091-0.08744 0.468 0.640869

cycleH-0.16573-0.07436 2.229 0.027999 *

cyclel-0.23225-0.08777 2.646 0.009418 **

cycleJ-0.34599-0.09252 3.740 0.000303 ***

Signif. codes: 0 ‘***’ 0.001 ‘**’ 0.01 ‘*’ 0.05 ‘.’ 0.1 ‘‘1

(Dispersion parameter for binomial family taken to be 1.000021)

Number of Fisher Scoring iterations: 4

“

### NHANES-GPT

The summary of your logistic regression model, which examines the trends in obesity across different NHANES cycles without controlling for confounders, provides several useful insights:

### Model Coefficients

- The coefficients (Estimates) represent the log odds of the prevalence of obesity for each cycle compared to the reference cycle (assumed to be the earliest cycle, “2005-2006”).
- Positive coefficients indicate higher odds of obesity in that cycle compared to the reference, while negative coefficients indicate lower odds.

### Statistical Significance

- The `‘Pr(> It I)’` values are the p-values for each coefficient. They indicate whether the differences in obesity prevalence across cycles are statistically significant.
- Coefficients with p-values less than 0.05 (indicated by ‘*’, ‘**’,or ‘***’)are statistically significant. For example, cycles H, I, and J show significant increases in obesity prevalence compared to the reference cycle.

### Interpreting the Coefficients

- For cycles H, I, and J, the positive coefficients and their statistical significance suggest a significant increase in the prevalence of obesity in these cycles compared to the baseline cycle.
- The magnitude of the coefficient indicates the size of this effect. For instance, the coefficient for cycle J is 0.34599, which is the largest among the cycles, suggesting a more substantial increase in the prevalence of obesity in this cycle.

### Model Fit and Convergence

- The model has successfully converged as indicated by the ‘Number of Fisher Scoring iterations’.
- The dispersion parameter is close to 1, which is expected for a binomial logistic regression.

### Next Steps

- **Incorporate Confounders:** To get a more accurate understanding, you should include potential confounders (such as age, gender, race/ethnicity, socioeconomic status, and smoking status) in your model. This will help to adjust for factors that might influence the observed trends.
- **Re-Run the Model:** After including confounders, re-run the model and compare the results. Note any changes in the coefficients and their significance, as this will give you insight into how these variables might be affecting the relationship between the NHANES cycle and obesity prevalence.
- **Interpretation:** Carefully interpret the adjusted model in the context of the added variables. This will provide a more nuanced understanding of the trends in obesity prevalence across NHANES cycles.

Incorporating confounders and analyzing the adjusted model will give you a more comprehensive picture of the trends in obesity prevalence and the factors influencing these trends.

### You

When I incorporate confounding variables, this is the model summary. But the cycleE, cycleF, and cycleG are missing. Where did they go? And what is the interpretation of these model results?

“Call:

svyglm(formula = obesity ∼ cycle+ RIDAGEYR + RIAGENDR + RIDRETH3 + DMDEDUC2 + INDHHIN2 + SMQ020, design= nhanes_design, family= binomial())

Survey design:

svydesign(ids = ∼ SDMVPSU, strata= ∼ SDMVSTRA, weights= -WTMEC2YR, data= all_data, nest= TRUE)

Coefficients:

Estimate Std. Error t value Pr(>ltl)

(Intercept) -0.186464 0.123227 -1.513 0.136175

cycleH 0.119078 0.066879 1.780 0.080729.

cyclel 0.220441 0.078756 2.799 0.007133 **

cycleJ 0.330190 0.089881 3.674 0.000558 ***

RIDAGEYR 0.004044 0.001278 3.163 0.002584 **

RIAGENDR 0.123143 0.046402 2.654 0.010482 *

RIDRETH3 -0.103214 0.016289 -6.337 5.25e-08 ***

DMDEDUC2 -0.105894 0.018496 -5.725 4.94e-07 ***

INDHHIN2 -0.004746 0.001496 -3.172 0.002521 **

SMQ020 -0.025179 0.044175 -0.570 0.571095

---

Signif. codes: 0 ‘***’ 0.001 ‘**’ 0.01 ‘*’ 0.05 ‘.’ 0.1 ‘‘1

(Dispersion parameter for binomial family taken to be 1.141418)

Number of Fisher Scoring iterations: 4”

### NHANES-GPT

The absence of `‘cycleE’, ‘cycleF’`, and `‘cycleG’` in your model summary suggests that these categories of the `‘cycle’` variable have been automatically omitted by R due to multicollinearity or because they are perfectly collinear with the intercept and other cycles. In regression models in R, one level of each categorical variable is used as a reference category and is not shown in the output. The coefficients of other categories represent the effect relative to this reference category.

### Interpretation of the Model Results

#### Coefficients of **‘cycle’** Variable

- The coefficients for `‘cycleH’`, `‘cycle’`, and `‘cycleJ’` represent the log odds of obesity compared to the reference cycle (which seems to be `‘cycleD’` or `‘2005-2006’` in your case).
- These coefficients are positive and (for `‘cycle’` and `‘cycleJ’`) statistically significant, indicating a higher prevalence of obesity in these cycles compared to the reference cycle.

#### Coefficients of Confounders

- `‘RIDAGEYR’` (Age): The positive coefficient suggests that the odds of obesity increase with age.
- `‘RIAGENDR’` (Gender): This coefficient indicates the difference in the odds of obesity between genders.
- `‘RIDRETH3’` (Race/Ethnicity): The negative coefficient suggests lower odds of obesity in certain racial/ethnic groups compared to the reference group.
- `‘DMDEDUC2’` (Education Level): The negative coefficient implies that higher education levels are associated with lower odds of obesity.
- `‘INDHHIN2’` (Income Level): The negative coefficient indicates that higher income levels are associated with lower odds of obesity.
- `‘SMQ020’` (Smoking Status): The negative coefficient suggests a relationship between smoking and obesity, but it’s not statistically significant in your model.

#### Significance Levels

- The significance levels (indicated by stars) show how statistically significant each coefficient is. For example, the significant positive coefficients for `‘cycle’` and `‘cycleJ’` suggest a meaningful increase in obesity in these cycles compared to the reference cycle.

#### Model Fit

- The dispersion parameter is slightly above 1, which is still reasonable for a binomial model.
- The model has converged successfully as indicated by the number of Fisher Scoring iterations.

## Conclusion

This model provides insights into how the prevalence of obesity has changed over different NHANES cycles when adjusting for various demographic and lifestyle factors. It suggests that certain cycles have significantly higher odds of obesity compared to the reference cycle, and highlights the role of age, gender, race/ethnicity, education, and income in influencing obesity prevalence. The absence of statistical significance for smoking status suggests that, in this model, smoking is not significantly associated with obesity after controlling for other factors.

